# Device-quantified vigorous intermittent lifestyle physical activity and risk of incident depression and anxiety among non-exercising adults

**DOI:** 10.64898/2026.05.18.26353464

**Authors:** Xunyin Zhang, Keyi Si, Matthew Ahmadi, Nuo Chen, Mark Hamer, John Mitchell, Nicholas Koemel, Mengzhuo Qiu, Xuemei Wang, Jiahao Min, Emmanuel Stamatakis, Zhi Cao, Chenjie Xu

## Abstract

**Background:** Physical activity is a well-established modifiable risk factor of depression and anxiety. However, whether vigorous intermittent lifestyle physical activity (VILPA)—short, sporadic bouts embedded in daily life—confers mental health benefits remains unclear. We aimed to examine the associations of accelerometer-measured VILPA with risks of incident depression and anxiety among non-exercising adults.

**Methods:** This prospective cohort study included 19,962 non-exercising adults (mean age 62.3 years) from the UK Biobank, free of depression and anxiety at baseline (2013–2015), with 7-day wrist-worn accelerometry data. Cox proportional hazards models and restricted cubic splines were used to examine associations between average daily duration of VILPA bouts (up to 1 or 2 minutes) and these outcomes.

**Findings:** Over an average follow-up of 7.8 years, 469 participants developed depression and 536 developed anxiety. Approximately 94.6% of participants engaged in VILPA bouts lasting up to 1 minute. Daily VILPA duration exhibited L-shaped associations with both depression and anxiety. Compared with participants who accumulated no VILPA, the whole sample median daily VILPA duration (lasting up to 1 minute) of 4.1 minutes was associated with a hazard ratio of 0.70 (95% confidence interval [CI]: 0.56-0.88) for depression and 0.79 (95% CI: 0.64-0.97) for anxiety. Findings were similar for VILPA bouts lasting up to 2 minutes.

**Interpretation:** Among non-exercisers, even small amounts of VILPA are associated with substantially lower risks of depression and anxiety, highlighting the potential of high intensity incidental physical activity as a feasible strategy for preventing depression and anxiety, particularly among individuals unable or unwilling to engage in structured exercise.

**Research in context:** *Evidence before this study:* We searched PubMed and Web of Science, without language restrictions, using the terms (physical activity OR vigorous intensity OR VILPA) AND (depression OR anxiety OR mental health). Most previous studies on physical activity and mental health have focused on structured leisure-time activity, usually accumulated in bouts of at least 10–15 minutes, and have relied mainly on self-reported physical activity. Few studies have investigated short bouts of vigorous activity embedded within daily living. Research on vigorous intermittent lifestyle physical activity (VILPA) is emerging, but has not specifically focused on mental health outcomes. Thus, evidence directly addressing whether brief intermittent vigorous activity undertaken as part of everyday life is associated with depression, anxiety, or broader mental health remains scarce.

*Added value of this study:* This study provides prospective evidence that device-measured daily VILPA is associated with lower risks of incident depression and anxiety. We found Daily VILPA duration exhibited L-shaped associations with both depression and anxiety, with small amounts of VILPA were associated with substantially lower risk of depression and anxiety. Compared with no VILPA, the median daily VILPA duration of 4.1 minutes was associated with a 30% lower risk of depression and a 21% lower risk of anxiety.

*Implications of all the available evidence:* VILPA might represent a feasible and accessible alternative for adults who are unwilling or unable to engage in structured leisure-time exercise, for whom barriers to participation can be substantial. The widespread use of consumer wearable devices also creates new opportunities to capture brief bursts of non-exercise physical activity accumulated across the day. Together, the available evidence and our findings highlight the potential public health importance of brief intermittent vigorous physical activity undertaken as part of daily living, and may help inform future intervention design, target setting, and monitoring.

## Introduction

Depression and anxiety are the most prevalent mental disorders worldwide, affecting 279.6 million and 301.4 million individuals in 2019^1^ and represent the second and sixth leading causes of years lived with disability in 2021, respectively^2^. Physical activity (PA) has been widely recognized as protective against depression and anxiety^3,4^, with evidence suggesting that vigorous physical activity (VPA) may confer greater benefits than moderate PA (MPA). Over 75 min/week of VPA has been estimated to account for 18.9% of potentially preventable affective disorder cases, compared with 2.6% for 150–300 min/week of moderate PA^5^. However, despite the well-established benefits of PA, approximately 31% of adults globally remained physically inactive in 2022^6^. Lack of time was the most commonly reported cited barrier, rendering conventional, structured PA recommendations impractical for many individuals^7^. Non-exercising adults constitute a large population with limited access to exercise-based interventions and a higher risk of depression and anxiety^3,8^. This highlights the need for alternative forms of PA that are accessible, time-efficient, and embedded into daily life. Brief lifestyle-embedded activities may overcome time barriers without requiring dedicated exercise sessions or special equipment, and thus could serve as a feasible and sustainable source of physical activity for non-exercising adults.

Advances in wearable technology have enabled the objective capture of short, sporadic bouts of VPA in free-living settings that were previously difficult to quantify. Emerging evidence suggests that transient bursts of vigorous exertion may confer health benefits beyond those attributable to total PA volume alone^9,10^. In parallel, the 2018 US PA Guidelines and 2020 WHO Guidelines removed the previous 10-minute minimum bout requirement, recognizing that PA of any duration contributes to health^11,12^. Together, these methodological and conceptual advances have reshaped how PA is characterized and have given rise to the concept of vigorous intermittent lifestyle PA (VILPA). VILPA refers to very short (typically 1–2 minutes), high-intensity movements that occur naturally in daily life—such as climbing stairs, brisk uphill walking, or carrying heavy loads^13^. Unlike structured exercise, VILPA requires no dedicated time, facilities, or equipment, making it particularly feasible for inactive adults and conceptually resonating with the notion of ‘exercise snacks’, as it reduces traditional barriers by emphasizing movements embedded in daily life^14^. Our previous studies have linked VILPA to lower risk of all-cause mortality, cancer incidence, and cardiovascular events^15–17^. However, its potential implications for mental disorders have not yet been investigated.

The aim of this study was to explore the associations of daily VILPA duration with risks of depression and anxiety in a large wearable-based cohort including participants who reported no leisure-time exercise and measured PA by accelerometers. This will bridge existing gaps in understanding the implications of brief, intermittent vigorous activity for mental disorders.

## Methods

### Study design and participants

The UK Biobank is an ongoing population-based prospective cohort that recruited more than 500,000 adults aged 37–73 years between 2006 and 2010 (baseline), when information on demographics, lifestyle, medication use, and mental health was collected using structured touch-screen questionnaires. All participants provided written informed consent for longitudinal linkage to hospital admission records. Ethical oversight was ensured by the NHS National Research Ethics Service (11/NW/0382).

Between 2013 and 2015, 236,462 participants with valid email addresses were invited to participate in an accelerometer sub-study to monitor PA and sleep. A total of 103,684 participants accepted the invitation and were sent an Axivity AX3 device (Newcastle upon Tyne, UK) to be worn on the dominant wrist continuously for 24 hours per day over seven consecutive days, with a sampling frequency of 100 Hz. A monitoring day was considered valid if wear time exceeded 16 hours. Participants were eligible for the accelerometer analyses if they contributed at least three valid days, including at least one weekend day^18^.

Our study focused on non-exercisers who did not report any leisure-time exercise (such as swimming or cycling) or more than one recreational walk per week, as assessed by a dedicated leisure-time physical activity questionnaire on activity type, duration, and frequency (**Supplementary Table 1**). We also excluded participants with pre-existing depression or anxiety at the time of accelerometry, identified through linked primary care data, hospital inpatient records, and self-reported medical conditions to reduce the likelihood of missed diagnoses. Additional exclusions were applied for participants who reported an inability to walk, had insufficient valid accelerometer wear days, or had missing covariate information. Finally, 19,962 participants were included in the primary analysis.

### Physical activity assessment

We applied a validated two-stage, machine learning-based random forest classifier to derive PA metrics from the accelerometer data^15,19^. In the first stage, activities were categorized into sedentary behavior, standing utilitarian movements, walking-related activities, and running or other high-energy activities. In the second stage, walking-related activities were further classified into light (<3 metabolic equivalents [METs]), moderate (3–<6 METs), and vigorous (≥6 METs) intensity levels. Vigorous ambulatory-related activities and running or other high-energy activities were identified as candidates for VILPA. Further details regarding the methods and accuracy of the classification scheme are provided in the **Supplementary methods.**

In this study, VILPA was operationalized as short, intermittent bouts lasting ≤2 minutes, consistent with evidence indicating that most real-world VILPA episodes reach vigorous physiological intensity within 1-2 minutes^20^. We performed detailed analyses on bouts lasting up to 1 minute and presented indicative findings for those up to 2 minutes.

### Ascertainment of depression and anxiety

Information on the date of depression/anxiety diagnosis was obtained from the ‘first occurrences’ dataset in the UK Biobank (category ID 1712), a multisource dataset that integrates information from primary care data, hospital inpatient records, death registers, and self-reported medical conditions. Diagnoses from these sources were mapped to three-character International Classification of Disease (ICD) codes. Specifically, depression was defined as depressive episode and recurrent depressive disorder (ICD-10 codes: F32-F33), while anxiety was defined as phobic anxiety disorders and other anxiety disorders (ICD-10 codes: F40-F41). Participants were followed from the date of accelerometer wear completion until the first diagnosis of the relevant outcome, death, or the end of follow-up (1 Mar 2023 for depression and 26 Oct 2022 for anxiety), whichever occurred first.

### Covariates

Covariates included baseline age (years, continuous), sex (male or female), ethnicity (White or non-White), body mass index (BMI, continuous), educational attainment (college/university degree or other degrees), Townsend deprivation index (TDI, continuous), smoking status (never, former, or current), drinking status (never, former, within guidelines, or above guidelines), dietary pattern (healthy or unhealthy)^21^, and self-reported medication use diabetes, hypertension, or hypercholesterolemia (yes or no). Additional covariates included accelerometry-estimated sleep duration (hours/day, continuous), PA energy expenditure volume from non-VILPA (continuous), and daily duration of VPA bouts lasting >1 minute (for analyses of VILPA bouts ≤1 minute) or >2 minutes (for analyses of VILPA bouts ≤2 minutes)^15^. Details of covariate definitions are presented in **Supplementary Table 2**.

### Statistical analyses

In our study, the VILPA values were capped at the 97.5 percentile to reduce the influence of sparse data or extreme outliers^15–18^. Baseline characteristics of the participants were summarized as means and standard deviations (SDs) for continuous variables and as numbers and percentages for categorical variables. Cox proportional hazards models were performed to estimate the hazard ratios (HRs) and 95% confidence intervals (CIs) for depression and anxiety associated with daily duration of VILPA bouts lasting up to 1 minute and up to 2 minutes^15^. Furthermore, we used restricted cubic splines to examine the dose-response associations of VILPA and outcomes^22^. Departure from linearity was assessed by a Wald test. Proportional hazards assumptions were tested using Schoenfeld residuals in the models with all these outcomes, and no violations were observed (all *P* > 0.05). To provide conservative point estimates we calculated the ‘minimal dose’, defined as VILPA duration associated with 50% of the maximal risk reduction^15,19,23,24^. We also reported HRs and 95% CIs corresponding to the median duration VILPA values.

Stratified analyses were conducted by age (<65 and ≥65 years) and sex (female and male) and multiplicative interactions were tested to tested for the possibility of age- and sex-differences. A series of sensitivity analyses were performed to test the robustness of the findings. First, we included the accelerometer wear season as an additional covariate to account for potential variance in exposure and to improve estimate precision. Second, to minimize potential reverse causation and account for prodromal mental health conditions, we excluded participants who developed depression or anxiety within the first 2 years of follow-up, those with baseline depressive or anxiety symptoms as assessed by the corresponding subscales of the Patient Health Questionnaire-4 (PHQ-4), and individuals reporting no daily VILPA or poor self-rated health, and participants using antidepressant or antipsychotic medications at accelerometry baseline. Third, given the potential impact of cancer and cardiovascular disease (CVD) on mental health outcomes, we replicated the analysis after excluding participants with prevalent cancer or CVD at the time of accelerometry assessment. Fourth, we used alternative reference values of VILPA in the dose-response association. Fifth, competing risk analyses were performed using Fine–Gray subdistribution hazard models, treating death from other causes as a competing event.

We performed all analyses using R statistical software (version 4.4.1) with the *rms* package (version 7.0.0) and the *survival* package (version 3.8.3).

## Results

### Participant characteristics

Baseline characteristics of participants stratified by daily VILPA duration are presented in **Table 1**. The mean age was 62.3 years old (SD: 7.6 years) and 55.6% were female. Over a median follow-up of 7.8 years, 469 depression cases and 536 anxiety cases were recorded. Approximately 94.6% of participants engaged in some VILPA. Participants with higher VILPA duration were less likely to be current smokers and reported lower medication use.

**Table 1.** Baseline characteristics of participants by daily VILPA duration.

| Characteristics | Total, n | <1 min | 1 to <3 min | 3 to <5 min | ≥5 to <10 min | ≥10min |
| --- | --- | --- | --- | --- | --- | --- |
| **Sample size, n** | 19,962 | 3,860 | 4,754 | 2,907 | 4,040 | 4,401 |
| **Follow up of depression, years** | 8.09 (1.24) | 7.94 (1.53) | 8.10 (1.25) | 8.11 (1.23) | 8.14 (1.08) | 8.17 (1.07) |
| **Follow up of anxiety, years** | 7.77 (1.19) | 7.64 (1.42) | 7.77 (1.19) | 7.78 (1.17) | 7.80 (1.06) | 7.82 (1.06) |
| **Age, mean (SD)** | 62.31 (7.59) | 64.73 (7.00) | 63.25 (7.38) | 62.10 (7.43) | 61.41 (7.61) | 60.15 (7.66) |
| **BMI, mean (SD)** | 27.32 (4.84) | 28.87 (5.57) | 27.79 (4.97) | 27.17 (4.57) | 26.68 (4.39) | 26.14 (4.09) |
| **TDI, mean (SD)** | -1.56 (2.87) | -1.36 (2.98) | -1.55 (2.88) | -1.66 (2.84) | -1.67 (2.80) | -1.60 (2.85) |
| **Sex, n (%)** |  |  |  |  |  |  |
| Female | 11,105 (55.63) | 2,599 (67.33) | 2,907 (61.15) | 1,637 (56.31) | 2,081 (51.51) | 1,881 (42.74) |
| Male | 8,857 (44.37) | 1,261 (32.67) | 1,847 (38.85) | 1,270 (43.69) | 1,959 (48.49) | 2,520 (57.26) |
| **Ethnicity, n (%)** |  |  |  |  |  |  |
| White | 19,248 (96.42) | 3,756 (97.31) | 4,601 (96.78) | 2,800 (96.32) | 3,898 (96.49) | 4,193 (95.27) |
| Non-White | 714 (3.58) | 104 (2.69) | 153 (3.22) | 107 (3.68) | 142 (3.51) | 208 (4.73) |
| **Smoking history, n (%)** |  |  |  |  |  |  |
| Current | 1,708 (8.56) | 414 (10.73) | 406 (8.54) | 234 (8.05) | 320 (7.92) | 334 (7.59) |
| Never | 11,388 (57.05) | 2,092 (54.20) | 2,675 (56.27) | 1,682 (57.86) | 2,322 (57.48) | 2,617 (59.46) |
| Former | 6,866 (34.40) | 1,354 (35.08) | 1,673 (35.19) | 991 (34.09) | 1,398 (34.60) | 1,450 (32.95) |
| **Alcohol consumption, n (%)** |  |  |  |  |  |  |
| Never | 746 (3.74) | 160 (4.15) | 180 (3.79) | 99 (3.41) | 144 (3.56) | 163 (3.70) |
| Former | 572 (2.87) | 128 (3.32) | 150 (3.16) | 95 (3.27) | 94 (2.33) | 105 (2.39) |
| Within guidelines | 11,893 (59.58) | 2,434 (63.06) | 2,881 (60.60) | 1,718 (59.10) | 2,330 (57.67) | 2,530 (57.49) |
| Above guidelines | 6,751 (33.82) | 1,138 (29.48) | 1,543 (32.46) | 995 (34.23) | 1,472 (36.44) | 1,603 (36.42) |
| **Education, n (%)** |  |  |  |  |  |  |
| College or above | 7,648 (38.31) | 1,472 (38.13) | 1,834 (38.58) | 1,113 (38.29) | 1,561 (38.64) | 1,668 (37.90) |
| Below college | 12,314 (61.69) | 2,388 (61.87) | 2,920 (61.42) | 1,794 (61.71) | 2,479 (61.36) | 2,733 (62.10) |
| **Medication use, n (%)** |  |  |  |  |  |  |
| Yes | 4,875 (24.42) | 1,288 (33.37) | 1,248 (26.25) | 687 (23.63) | 865 (21.41) | 787 (17.88) |
| No | 15,087 (75.58) | 2,572 (66.63) | 3,506 (73.75) | 2,220 (76.37) | 3,175 (78.59) | 3,614 (82.12) |
| **Dietary pattern, n (%)** |  |  |  |  |  |  |
| Healthy | 9,937 (49.78) | 1,925 (49.87) | 2,451 (51.56) | 1,429 (49.16) | 1,999 (49.48) | 2,133 (48.47) |
| Unhealthy | 10,025 (50.22) | 1,935 (50.13) | 2,303 (48.44) | 1,478 (50.84) | 2,041 (50.52) | 2,268 (51.53) |
| **Sleep (hours per day), mean (SD)** | 7.24 (1.03) | 7.20 (1.11) | 7.26 (1.05) | 7.28 (1.03) | 7.25 (0.96) | 7.22 (0.99) |

*VILPA*, vigorous intermittent lifestyle physical activity; *SD*, standard deviation; *BMI*, body mass index; *TDI*, Townsend deprivation index.
Alcohol consumption: above guidelines is >14 units per week, where 1 unit = 8g of ethanol. Dietary score: based on the frequency of consumption of fruits, vegetables, fish, processed meat, unprocessed red meat, whole grains, and refined grains.

### Association of VILPA with incident depression

An L-shaped dose-response relationship was observed between daily VILPA duration and depression risk, with the steepest dose-response as daily VILPA increased from 0 to 6 minutes (*P* for non-linearity = 0.033; **Figure 1A**). The minimal daily dose for VILPA duration from bouts lasting up to 1 minute was 2.6 minutes per day, corresponding to a 21% significantly lower risk of depression (**Table 2**). The whole sample median daily VILPA duration of 4.1 minutes was associated with an HR of 0.70 (95% CI 0.56, 0.88). Results for bouts lasting up to 2 minutes were consistent with the main results in terms of the dose-response curve shapes, minimal dose estimates, and the magnitude of associations at the median daily duration (**Figure 1C**, **Table 2**).

**Figure 1.**
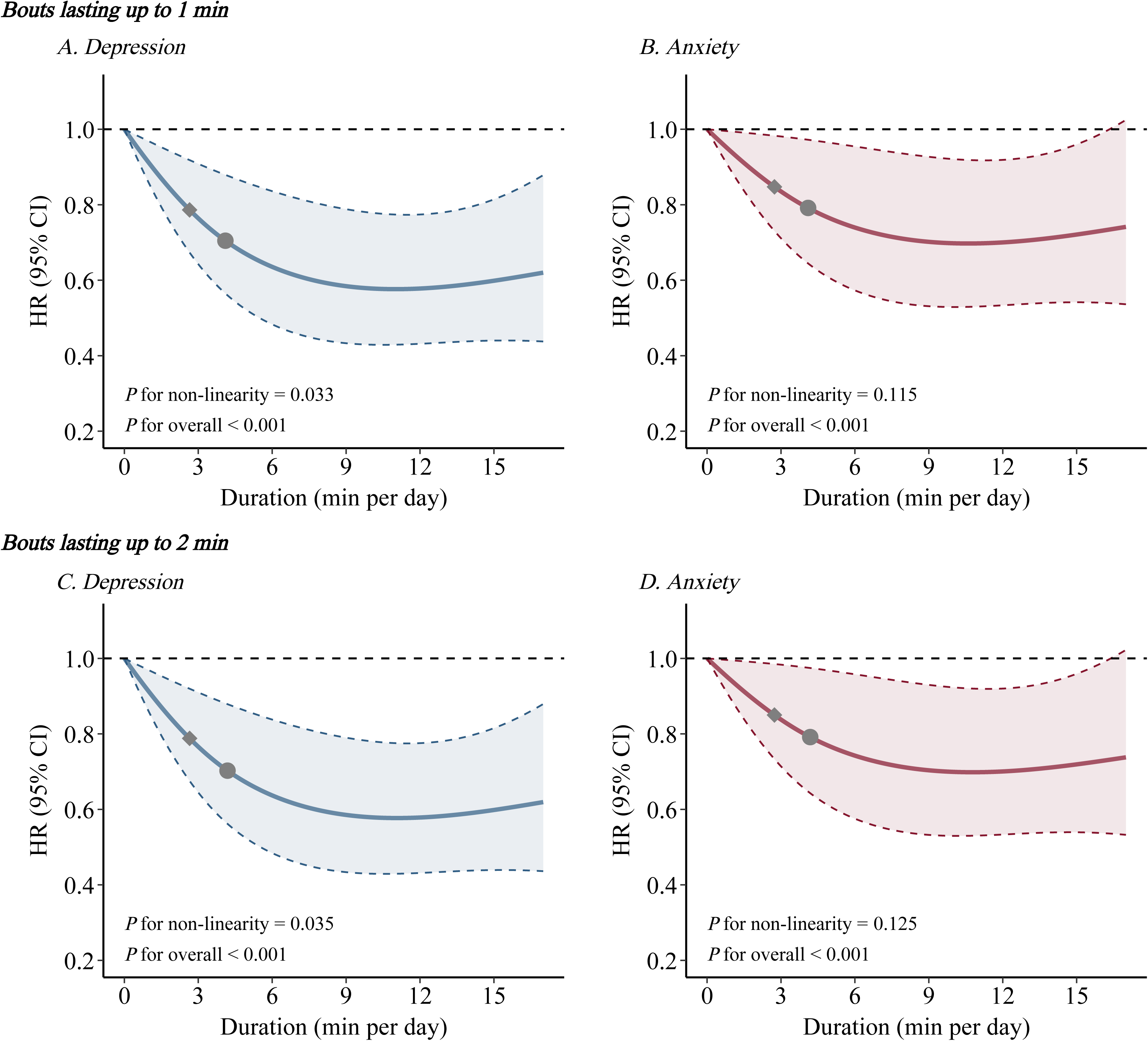
Association between daily duration of VILPA and mental disorders. *VILPA,* Vigorous intermittent lifestyle physical activity; *HRs*, hazard ratios; *CI,* confidence interval. A, B, Dose–response curves showing depression (A) and anxiety (B) HRs associated with increasing daily duration of VILPA, for bouts of VILPA up to 1 min duration. C, D, Dose–response curves showing depression (C) and anxiety (D) HRs associated with increasing daily duration of VILPA, for bouts of VILPA up to 2 min. Restricted cubic splines were constructed with three knots located at the 10th, 50th, and 90th percentiles of each exposure. Diamond, minimal dose, as indicated by the ED_50_ statistic which estimates the daily duration of VILPA associated with 50% of the maximal risk reduction. Circle, HR associated with the median VILPA value. Analyses were adjusted for age, sex, ethnicity, BMI, educational attainment, TDI, smoking history, drinking status, dietary pattern, and self-reported medication use (diabetes, hypertension, or hypercholesterolemia). Additional covariates included accelerometer estimated sleep duration, PA energy expenditure volume from non-VILPA, and daily duration of VPA bouts lasting >1 minute (for analyses of VILPA bouts ≤1 minute) or >2 minutes (for analyses of VILPA bouts ≤2 minutes). The shaded region demarcated by dashed lines represents the 95% CI. The solid line that lies within the shaded region represents the HR.

**Table 2.** Hazard ratios associated with the minimum dose, median VILPA values and the lowest estimated hazard among non-exercisers for up to 1 and up to 2 minutes bouts.

| **Outcome** | **Dose** | **HR (95% CI)** |
| --- | --- | --- |
| **Depression** |  |  |
| up to 1 min bout minimum dose | 2.6 | 0.79 (0.67, 0.92) |
| up to 1 min bout median VILPA dose | 4.1 | 0.70 (0.56, 0.88) |
| up to 1 min bout lowest estimated hazard dose | 11.0 | 0.58 (0.43, 0.77) |
| up to 2 min bout minimum dose | 2.6 | 0.79 (0.68, 0.92) |
| up to 2 min bout median VILPA dose | 4.2 | 0.70 (0.56, 0.88) |
| up to 2 min bout lowest estimated hazard dose | 11.0 | 0.58 (0.43, 0.78) |
| **Anxiety** |  |  |
| up to 1 min bout minimum dose | 2.7 | 0.85 (0.73, 0.98) |
| up to 1 min bout median VILPA dose | 4.1 | 0.79 (0.64, 0.97) |
| up to 1 min bout lowest estimated hazard dose | 10.6 | 0.70 (0.53, 0.92) |
| up to 2 min bout minimum dose | 2.7 | 0.85 (0.73, 0.99) |
| up to 2 min bout median VILPA dose | 4.2 | 0.79 (0.64, 0.97) |
| up to 2 min bout lowest estimated hazard dose | 10.8 | 0.70 (0.53, 0.92) |

*VILPA*, vigorous intermittent lifestyle physical activity; *HR*, Hazard ration; *CI*, confidence interval.
Minimal dose: defined as the duration of VILPA associated with 50% of the maximal risk reduction. The VILPA duration median values were calculated in the sample excluding participants with zero VILPA. Analyses adjusted for age, sex, ethnicity, BMI, educational attainment, TDI, smoking history, drinking status, dietary pattern, and self-reported medication use (diabetes, hypertension, or hypercholesterolemia). Additional covariates included accelerometer estimated sleep duration, PA energy expenditure volume from non-VILPA, and daily duration of VPA bouts lasting >1 minute (for analyses of VILPA bouts ≤1 minute) or >2 minutes (for analyses of VILPA bouts ≤2 minutes).

### Association of VILPA with anxiety disorder

Dose-response analyses revealed an L-shaped association of daily VILPA duration with anxiety (*P* for non-linearity = 0.115, **Figure 1B**). The minimal daily dose for VILPA duration from bouts lasting up to 1 minute was 2.7 minutes per day, corresponding to an HR of 0.85 (95% CI 0.73, 0.98). The whole sample median daily VILPA duration of 4.1 minutes was associated with an HR of 0.79 (95% CI 0.64, 0.97). Findings for bouts lasting up to 2 minutes were similar, with comparable dose-response curves and other metrics. (**Figure 1D**, **Table 2**).

### Stratified and sensitivity analyses

In stratified analyses by age, we found that modest amounts of daily VILPA were associated with a lower risk of depression among older participants (HR 0.57, 95% CI 0.41–0.79). In younger participants, the association was weaker and not statistically significant (HR 0.84, 95% CI 0.62–1.14), with no evidence of effect modification by age (*P* for interaction = 0.134) (**Figure 2, Supplementary table 3**). When the analyses were stratified by sex, no significant multiplicative interactions were observed between daily VILPA duration and the risk of depression or anxiety (all *P* for interaction > 0.05; **Figure 2-3, Supplementary table 3-4**). These findings indicate that the associations between VILPA and mental health outcomes were broadly consistent across sex and age groups.

**Figure 2.**
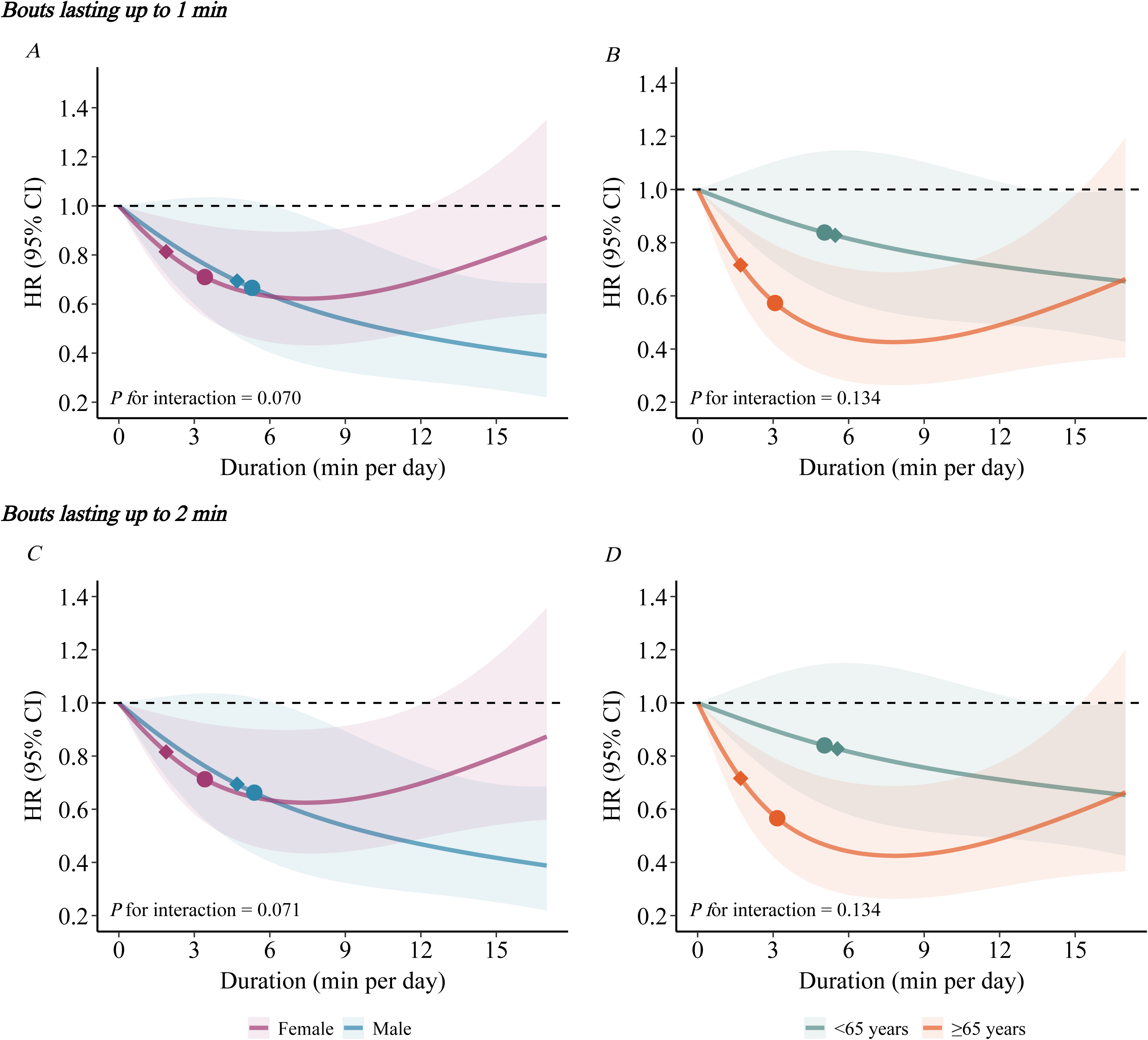
Association between VILPA duration and depression stratified by age and sex. *VILPA,* Vigorous intermittent lifestyle physical activity; *HRs*, hazard ratios; *CI,* confidence interval. A, B, Dose–response curves showing depression HRs associated with increasing daily duration of VILPA stratified by sex (A, female and male) and age (B, <65 and ≥ 65 years), for bouts of VILPA up to 1 min. C, D, Dose–response curves showing depression HRs associated with increasing daily duration of VILPA stratified by sex (C, female and male) and age (D, <65 and ≥ 65 years), for bouts of VILPA up to 2 min. Analyses adjusted for age, sex, ethnicity, BMI, educational attainment, TDI, smoking history, drinking status, dietary pattern, and self-reported medication use (diabetes, hypertension, or hypercholesterolemia). Additional covariates included accelerometer estimated sleep duration, PA energy expenditure volume from non-VILPA, and daily duration of VPA bouts lasting >1 minute (for analyses of VILPA bouts ≤1 minute) or >2 minutes (for analyses of VILPA bouts ≤2 minutes). Restricted cubic splines were constructed with three knots located at the 10th, 50th, and 90th percentiles of each exposure. Stratified factors were not adjusted for in the models. Diamond, minimal dose, as indicated by the ED_50_ statistic which estimates the daily duration of VILPA associated with 50% of the optimal risk reduction. Circle, HR associated with the median VILPA value (see Supplementary Table 3 for the list of values). The shaded region demarcated by dashed lines represents the 95% CI. The solid line that lies within the shaded region represents the HR. Wald tests were used to evaluate the significance of interaction terms.

**Figure 3.**
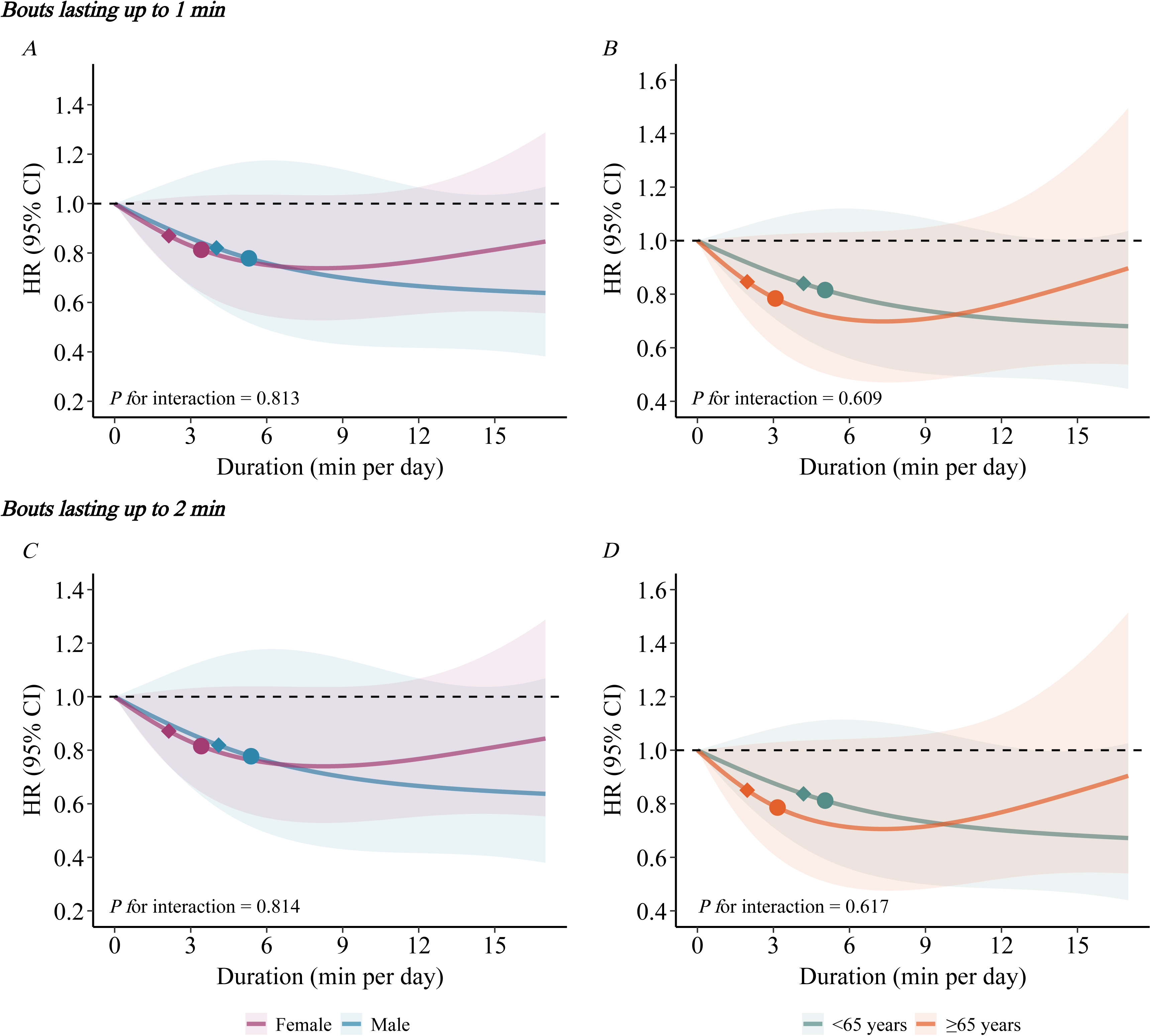
Association between VILPA duration and anxiety stratified by age and sex. *VILPA,* Vigorous intermittent lifestyle physical activity; *HRs*, hazard ratios; *CI,* confidence interval. A, B, Dose–response curves showing anxiety HRs associated with increasing daily duration of VILPA stratified by sex (A, female and male) and age (B, <65 and ≥ 65 years), for bouts of VILPA up to 1 min. C, D, Dose–response curves showing anxiety HRs associated with increasing daily duration of VILPA stratified by sex (C, female and male) and age (D, <65 and ≥ 65 years), for bouts of VILPA up to 2 min. Analyses adjusted for age, sex, ethnicity, BMI, educational attainment, TDI, smoking history, drinking status, dietary pattern, and self-reported medication use (diabetes, hypertension, or hypercholesterolemia). Additional covariates included accelerometer estimated sleep duration, PA energy expenditure volume from non-VILPA, and daily duration of VPA bouts lasting >1 minute (for analyses of VILPA bouts ≤1 minute) or >2 minutes (for analyses of VILPA bouts ≤2 minutes). Restricted cubic splines were constructed with three knots located at the 10th, 50th, and 90th percentiles of each exposure. Stratified factors were not adjusted for in the models. Diamond, minimal dose, as indicated by the ED_50_ statistic which estimates the daily duration of VILPA associated with 50% of the optimal risk reduction. Circle, HR associated with the median VILPA value (see Supplementary Table 4 for the list of values). The shaded region demarcated by dashed lines represents the 95% CI. The solid line that lies within the shaded region represents the HR. Wald tests were used to evaluate the significance of interaction terms.

The sensitivity analyses confirmed the robustness of the primary findings. Consistent results were observed after additional adjustment for accelerometer wear season (**Supplementary Figure 2)**, and upon excluding participants with prevalent cancer or CVD, those who experienced depression or anxiety events within the first 2 years of follow-up, or participants reporting poor health, or those using antidepressant or antipsychotic medications. (**Supplementary Figure 3-8**). For example, the median daily VILPA duration of 4.3 minutes was associated with an HR of 0.71 (95% CI 0.56, 0.91) for depression, and 0.79 (95% CI 0.64, 0.98) for anxiety after excluding participants with poor health. When excluding participants with depressive and anxiety symptoms, the dose-response curves were broadly consistent with the main analyses, although 95% CIs crossed unity at higher levels of daily VILPA duration (**Supplementary Figure 9**). Similarly, using alternative reference values of VILPA and treating death from other causes as a competing event did not appreciably change the results (**Supplementary Figure 10-11**).

## Discussion

In this large prospective study of non-exercising adults, we found that accelerometer-derived VILPA was associated with lower risks of depression and anxiety, following an approximately L-shaped dose-response pattern even when accumulated in very short and sporadic bouts. Relatively modest amounts of VILPA, such as the median duration of 4.1 minutes per day was associated with a 21% and 30% lower risk of anxiety and depression compared with no daily VILPA. These findings highlight the potential relevance of VILPA embedded in daily life.

Previous studies using both self-reported and accelerometer-derived measures have reported inverse, L-shaped associations between PA and the risks of depression and anxiety^3,25,26^. A meta-analysis of 49 prospective cohort studies showed that individuals with higher levels of self-reported PA had approximately 17% (95% CI, 12%, 21%) lower odds of developing depression^4^. Risk reductions of similar magnitude and comparable dose-response patterns have also been observed for accelerometer-measured MPA or VPA, whether accumulated through regular exercise or concentrated on weekends^5,27^. Together, these findings underpin current PA guidelines, which have traditionally emphasized planned MPA or VPA. Although the 2018 U.S. PA Guidelines removed the requirement that activity be accumulated in bouts of at least 10 minutes and WHO PA guidelines emphasized all activity counts regardless of domain or bout duration, acknowledging the potential contribution of lifestyle-based movement^11,12^. Nevertheless, the mental health benefits of brief, intermittent VPA embedded in daily life remain largely unexplored. This evidence gap is particularly relevant for physically inactive individuals, who comprise about one quarter to one third of the adult population^6,28^.

As a complementary alternative to structured exercise, VILPA focuses on brief, sporadic bouts of VPA embedded in daily life. Typical examples include stair climbing, brisk uphill walking, fast-paced commuting, and carrying heavy loads^13^. Unlike traditional MPA or VPA, which is usually accumulated through structured and sustained sessions, VILPA is incidental, requires no dedicated time or equipment, and is unconstrained by bout duration, potentially lowering the threshold for PA among individuals who are unwilling or unable to engage in regular exercise.

The approximately 30% reduction in depression risk associated with the median VILPA duration of 4.1 minutes per day is comparable to the 28% (95% CI: 19%, 36%) reduction associated with 17.5 marginal metabolic equivalent task hours per week (approximately 150 minutes per week of questionnaire-measured VPA)^3^. This apparent discrepancy in the volume of VPA required to achieve similar risk reductions may be partly explained by differences in PA measurement methods. Self-reported questionnaires typically capture recalled exercise bouts that include intermittent rest periods, whereas accelerometer-based approaches quantify the actual time spent at vigorous intensity. Additionally, achieving a comparable 30% reduction in depression risk required substantially less VILPA than device-measured MPA (141 minutes per week), as reported in our previous study^25^. However, the magnitude of risk reduction attainable at higher volumes differed across activity types. Both VILPA and MPA exhibited plateau effects, with maximal risk reductions of 38% at 11 minutes per day and 69% at 660 minutes per week, respectively. The apparent plateau for VILPA may largely reflect the sample distribution at higher levels of VILPA. In contrast, VPA showed a steep initial decline in risk (50%) up to approximately 50 minutes per week, followed by a more gradual decrease without clear evidence of a plateau^25^. These patterns suggest that VILPA may represent a time-efficient strategy for reducing depression risk among non-exercisers. Similar patterns were observed for anxiety.

Notably, based on our estimates, more than 94.6% of non-exercising adults accumulated some VILPA as part of their daily lives. However, approximately 40% of participants remained below the level associated with achieving half of the maximal risk reduction for depression and anxiety (about 2.6–2.7 minutes per day), and nearly 80% accumulated less than the duration associated with maximal risk reduction (about 10.6–11.0 minutes per day). This gap highlights the need for more effective promotion of non-structured activities like VILPA, as many individuals may not recognize such brief, vigorous lifestyle activities as meaningful-for-health PA. Therefore, interventions should aim to enhance situational awareness, enabling individuals recognize naturally occurring opportunities—such as walking faster during a commute or taking stairs—and intentionally amplify them, so that these behaviors can be gradually incorporated into daily routines as habitual responses to environmental cues. Qualitative research had shown that limited awareness of what constitutes VILPA and uncertainty about its health relevance represented key barriers to engagement, despite generally positive attitudes toward its feasibility^29^. Consistent with these findings, a recent mixed-methods study evaluated the promotion of VILPA via a mobile application (MovSnax) in a small sample of middle-aged adults and experts, finding VILPA to be acceptable and achievable but noting the need for clearer feedback and enhanced motivational support^30^. Future interventions could improve awareness through concise, engaging visual materials—such as short videos or infographics—illustrating VILPA’s potential benefits, complemented by community-or digitally-based activities to foster motivation and social support. Adaptable small goals could help individuals integrate VILPA into daily routines, while tracking or gamified elements may further support adherence^31,32^.

The underlying mechanisms linking VILPA with lower risk of depression and anxiety are likely complex and multifactorial, encompassing biological and psychosocial pathways. For example, PA may enhance brain health by stimulating neurogenesis and angiogenesis^33^, and alleviate psychiatric symptoms by modulating monoamines and neurotrophic factors (e.g., BDNF)^34^. Intermittent VPA may improve in cardiorespiratory fitness more efficiently than continuous exercise^35^, thus potentially strengthening psychological resilience and further reduce susceptibility to psychiatric disorders. Higher-intensity exercise has also been shown to improve inflammatory status and support neuroplasticity ^36^-critical factors in the pathogenesis of mental disorders. In addition to these biological mechanisms, psychosocial factors, such as enhanced self-esteem, self-concept, and self-efficacy, serve as key pathways through which PA may mitigate depressive and anxiety symptoms across the lifespan^37^. These complementary mechanisms provide a plausible explanation for the associations observed in this study and highlight potential targets for prevention.

### Strengths and limitations

The strengths of this study include prospective design, several sources for outcomes collection and meticulous control of diverse covariates. Moreover, we leveraged device-based PA measurements and a validated two-stage machine learning-based methods, ensuring objective and robust assessment^38^. Nonetheless, this study has several limitations. First, we cannot completely rule out reverse causation in observational studies, although the findings proved robust across relevant sensitivity analyses. Second, some VILPA (e.g., carrying a backpack) may not be fully captured by wrist-worn accelerometers, although such measurement error is likely to attenuate the ‘true’ associations with depressive and anxiety disorders through regression dilution bias. Third, most covariates and non-exerciser status were assessed at baseline (2006–2010), with a 5.5 year-gap to the accelerometry assessment. However, previous evidence indicates that these covariates are generally stable over time^39^ and non-exerciser status is a relatively stable factor (82–88% remained the non-exerciser status)^15^, suggesting that such a temporal gap is unlikely to impact the reliability of the current findings. Fourth, although PA patterns may vary with chronological age, accelerometry-measured PA has been shown to remain stable over time in adults (e.g., >90% of classification accuracy within one quartile over a period of 2–3 years)^40^.

## Conclusion

Our study found that VILPA was associated with a significantly lower risk of depression and anxiety in an L-shaped manner among non-exercising adults, suggesting that brief, vigorous lifestyle activities may constitute a scalable and low-barrier avenue for prevention of the common mental disorders alongside traditional exercise-based approaches. Recognizing VILPA as a meaningful form of PA may help inform PA guidance and population-level behavioral strategies for mental health.

## Acknowledgment

This study was conducted using the UK Biobank resource (Application NO.25813). We want to express our sincere thanks to the participants of the UK Biobank, and the members of the survey, development, and management teams of this project.

## Author contributions

ES, ZC and CX were involved in the conception, design, and interpretation of the results. MA was responsible for the processing of the wearables data and advised on statistical analyses. XZ, and KS wrote the first draft of the manuscript, and all authors edited, reviewed, and approved the final version of the manuscript. XZ and KS analyzed the data and interpretated the results. All authors have made critical revision of the manuscript for important intellectual content. CX is the guarantor of this work.

## Patient and public involvement

Patients and/or the public were not involved in the design, or conduct, or reporting or dissemination plans of this research.

## Patient consent for publication

Not applicable.

## Data availability statement

The data that we used in this study can be accessed by researchers on application. Requests for data access should be made directly to the UK Biobank.

## The use of AI and AI-assisted technologies in scientific writing

We did not use AI or AI-assisted technologies in the writing of this manuscript.

