## Supplementary Materials for "Device-quantified vigorous intermittent lifestyle physical activity and risk of incident depression and anxiety among non-exercising adults"

**Supplemental materials**

***Supplementary Table 1.* Questions to assess participation in leisure time physical activity**

***Supplementary Table 2.* Covariate definitions**

***Supplementary Table 3.* Hazard ratios of depression across age and sex associated with the minimum dose and median VILPA values among non-exercisers for up to 1 and up to 2 minutes bouts**

***Supplementary Table 4*. Hazard ratios of anxiety across age and sex associated with the minimum dose and median VILPA values among non-exercisers for up to 1 and up to 2 minutes bouts**

***Supplementary Figure 1.* Flowchart of participant selection**

***Supplementary Figure 2.* Dose-response association of VILPA duration with risks of depression and anxiety after further adjusting for accelerometer wear season**

***Supplementary Figure 3.* Dose-response association of VILPA duration with risks of depression and anxiety after excluding prevalent cancer at baseline**

***Supplementary Figure 4.* Dose-response association of VILPA duration with risks of depression and anxiety after excluding prevalent CVD at baseline**

***Supplementary Figure 5.* Dose-response association of VILPA duration with risks of depression and anxiety after excluding outcome diagnosed within 2 years since baseline**

***Supplementary Figure 6.* Dose-response association of VILPA duration with risks of depression and anxiety after excluding participants with poor health**

***Supplementary Figure 7.* Dose-response association of VILPA duration with risks of depression and anxiety after excluding participants using antidepressant or antipsychotic medications**

***Supplementary Figure 8.* Dose-response association of VILPA duration with risks of depression and anxiety after excluding participants with VILPA = 0 min /day**

***Supplementary Figure 9.* Dose-response association of VILPA duration with risks of depression and anxiety after excluding participants with baseline depressive and anxiety symptoms**

***Supplementary Figure 10.* Association of VILPA duration with risks of depression and anxiety using the minimal dose as the referent point**

***Supplementary Figure 11.* Association of VILPA duration with risks of depression and anxiety after treating death as a competing event**

***Supplementary Methods.* Wearable physical activity intensity and posture classification**

**Supplementary Table 1. Questions to assess participation in leisure time physical activity**

| Types of physical activity in last 4 weeks UK Biobank Field ID: 6164: | Walking for pleasure (not as a means of transport)  • Other exercises (e.g.: swimming, cycling, keep fit, bowling)  • Strenuous sports  • Light DIY (e.g.: pruning, watering the lawn)  •Heavy DIY (e.g.: weeding, lawn mowing, carpentry, digging)  • None of the above  • Prefer not to answer |
| --- | --- |
| Each time you did strenuous sports, about how long did you spend doing it? (UK Biobank field ID: 1001) | • Less than 15 minutes  • between 15 and 30 minutes  • between 30 minutes and 1 hour  • between 1 hour and 1.5 hours  • between 1.5 hours and 2 hours between 2 and 3 hours  • over 3 hours, do not know  • prefer not to answer |
| How many times in the last 4 weeks did you do strenuous sports? (UK Biobank field ID: 991) | • Once in the last 4 weeks  • 2-3 times in the last 4 weeks  • Once a week  • 2-3 times a week  • 4-5 times a week • Every day  • Do not know  • Prefer not to answer |
| Each time you did other exercises such as swimming, cycling, keep fit, about how long did you spend doing it? (UK Biobank field ID: 3647) | • Less than 15 minutes  • between 15 and 30 minutes  • between 30 minutes and 1 hour  • between 1 hour and 1.5 hours  • between 1.5 hours and 2 hours between 2 and 3 hours  • over 3 hours, do not know  • prefer not to answer |
| How many times in the last 4 weeks did you do other exercises such as swimming, cycling, keep fit? (UK Biobank field ID: 3637) | • Once in the last 4 weeks  • 2-3 times in the last 4 weeks  • Once a week  • 2-3 times a week  • 4-5 times a week  • Every day  • Do not know  • Prefer not to answer |
| How many times in the last 4 weeks did you go walking for pleasure? (UK Biobank field ID: 971) | • Once in the last 4 weeks  • 2-3 times in the last 4 weeks  • Once a week  • 2-3 times a week  • 4-5 times a week  • Every day  • Do not know  • Prefer not to answer |

**Supplementary Table 2. Covariate definitions**

| **Variable** | **Definition** | **UK Biobank field ID (if applicable)** |
| --- | --- | --- |
| Age | Continuous | 34, 52, accelerometer date-timestamp |
| Sex | Female/Male | 31 |
| Ethnicity | White or non-White | 21000 |
| VILPA duration from longer bouts | VILPA duration from bouts lasting over 1 or 2 minutes. e.g., for the analysis of bouts lasting up to 2 minutes in duration, the variable contained VILPA duration from bouts that lasted more than 2 minutes in duration | Derived from accelerometer data |
| Physical activity energy expenditure volume from non-VILPA | Physical activity energy expenditure volume from non-VILPA (kilojoules/kg/d, Continuous) | Derived from accelerometer data |
| Smoking status | Never, previous, current | 20116 |
| Alcohol consumption | Never, ex-drinker, within guidelines, above guidelines | 20117, 1558 |
| Sleep duration | Hours spent sleeping | Derived from accelerometer data (see Online Methods) |
| Dietary pattern | The dietary pattern based on the frequency of consumption of fruits, vegetables, fish, processed meat, unprocessed red meat, whole grains, and refined grains, categorized as unhealth (<4 score), and health (≧4 score) | 1289, 1299,1309, 1319, 1329, 1339, 1349, 1369, 1379, 1389, 1438, 1448, 1458, 1468 |
| Prevalent cancer | Identified by self-report and cancer registry | 20001, 100092 |
| Prevalent CVD | Identified by self-report and hospitalisation | 20002, 2000 |
| Education | below college/college or above | 6138 |
| Use of medication | Yes/No | 6177, 6153, 20003 |

**Supplementary Table 3. Hazard ratios of depression across age and sex associated with the minimum dose and median VILPA values among non-exercisers for up to 1 and up to 2 minutes bouts**

| **Subgroup** | **Dose** | **HR (95% CI)** |
| --- | --- | --- |
| **Female (n=11,105, event=303)** |  |  |
| up to 1 min bout minimum dose | 1.9 | 0.81 (0.70, 0.95) |
| up to 1 min bout median VILPA dose | 3.4 | 0.71 (0.55, 0.92) |
| up to 2 min bout minimum dose | 1.9 | 0.82 (0.70, 0.95) |
| up to 2 min bout median VILPA dose | 3.4 | 0.71 (0.55, 0.92) |
| **Male (n=8,857, event=166)** |  |  |
| up to 1 min bout minimum dose | 4.7 | 0.69 (0.47, 1.03) |
| up to 1 min bout median VILPA dose | 5.3 | 0.67 (0.43, 1.02) |
| up to 2 min bout minimum dose | 4.7 | 0.69 (0.47, 1.03) |
| up to 2 min bout median VILPA dose | 5.4 | 0.66 (0.43, 1.02) |
| **<65 years (n=11,432, event=289)** |  |  |
| up to 1 min bout minimum dose | 5.5 | 0.83 (0.59, 1.15) |
| up to 1 min bout median VILPA dose | 5.0 | 0.84 (0.62, 1.14) |
| up to 2 min bout minimum dose | 5.5 | 0.83 (0.60, 1.15) |
| up to 2 min bout median VILPA dose | 5.0 | 0.84 (0.62, 1.15) |
| **≥65 years (n=8,530, event=180)** |  |  |
| up to 1 min bout minimum dose | 1.7 | 0.72 (0.59, 0.87) |
| up to 1 min bout median VILPA dose | 3.1 | 0.57 (0.41, 0.79) |
| up to 2 min bout minimum dose | 1.7 | 0.72 (0.59, 0.87) |
| up to 2 min bout median VILPA dose | 3.2 | 0.57 (0.41, 0.79) |
| *VILPA*, vigorous intermittent lifestyle physical activity; *HR,* Hazard ration; *CI,* confidence interval.  Minimal dose: defined as the duration of VILPA associated with 50% of the optimal risk reduction. The VILPA duration median values were calculated in the sample excluding participants with zero VILPA. Analyses adjusted for age, sex, ethnicity, BMI, educational attainment, TDI, smoking history, drinking status, dietary pattern, and self-reported medication use (diabetes, hypertension, or hypercholesterolemia). Additional covariates included accelerometer estimated sleep duration, PA energy expenditure volume from non-VILPA, and daily duration of VPA bouts lasting >1 minute (for analyses of VILPA bouts ≤1 minute) or >2 minutes (for analyses of VILPA bouts ≤2 minutes). | | |

**Supplementary Table 4. Hazard ratios of anxiety across age and sex associated with the minimum dose and median VILPA values among non-exercisers for up to 1 and up to 2 minutes bouts**

| **Subgroup** | **Dose** | **HR (95% CI)** |
| --- | --- | --- |
| **Female (n=11,105, event=354)** |  |  |
| up to 1 min bout minimum dose | 2.1 | 0.87 (0.74, 1.02) |
| up to 1 min bout median VILPA dose | 3.4 | 0.81 (0.64, 1.03) |
| up to 2 min bout minimum dose | 2.1 | 0.87 (0.74, 1.02) |
| up to 2 min bout median VILPA dose | 3.4 | 0.82 (0.64, 1.03) |
| **Male (n=8,857, event=182)** |  |  |
| up to 1 min bout minimum dose | 4.0 | 0.82 (0.59, 1.15) |
| up to 1 min bout median VILPA dose | 5.3 | 0.78 (0.52,1.17) |
| up to 2 min bout minimum dose | 4.1 | 0.82 (0.58, 1.15) |
| up to 2 min bout median VILPA dose | 5.4 | 0.78 (0.52,1.17) |
| **<65 years (n=11,432, event=284)** |  |  |
| up to 1 min bout minimum dose | 4.2 | 0.84 (0.64, 1.11) |
| up to 1 min bout median VILPA dose | 5.0 | 0.81 (0.59, 1.12) |
| up to 2 min bout minimum dose | 4.2 | 0.84 (0.63, 1.10) |
| up to 2 min bout median VILPA dose | 5.0 | 0.81 (0.59, 1.12) |
| **≥65 years (n=8,530, event=252)** |  |  |
| up to 1 min bout minimum dose | 2.0 | 0.85 (0.70, 1.02) |
| up to 1 min bout median VILPA dose | 3.1 | 0.78 (0.60, 1.02) |
| up to 2 min bout minimum dose | 2.0 | 0.85 (0.71, 1.02) |
| up to 2 min bout median VILPA dose | 3.2 | 0.79 (0.60, 1.03) |
| *VILPA*, vigorous intermittent lifestyle physical activity; *HR,* Hazard ration; *CI,* confidence interval.  Minimal dose: defined as the duration of VILPA associated with 50% of the optimal risk reduction. The VILPA duration median values were calculated in the sample excluding participants with zero VILPA. Analyses adjusted for age, sex, ethnicity, BMI, educational attainment, TDI, smoking history, drinking status, dietary pattern, and self-reported medication use (diabetes, hypertension, or hypercholesterolemia). Additional covariates included accelerometer estimated sleep duration, PA energy expenditure volume from non-VILPA, and daily duration of VPA bouts lasting >1 minute (for analyses of VILPA bouts ≤1 minute) or >2 minutes (for analyses of VILPA bouts ≤2 minutes). | | |

**Supplementary Figure. 1 Flowchart of participant selection**

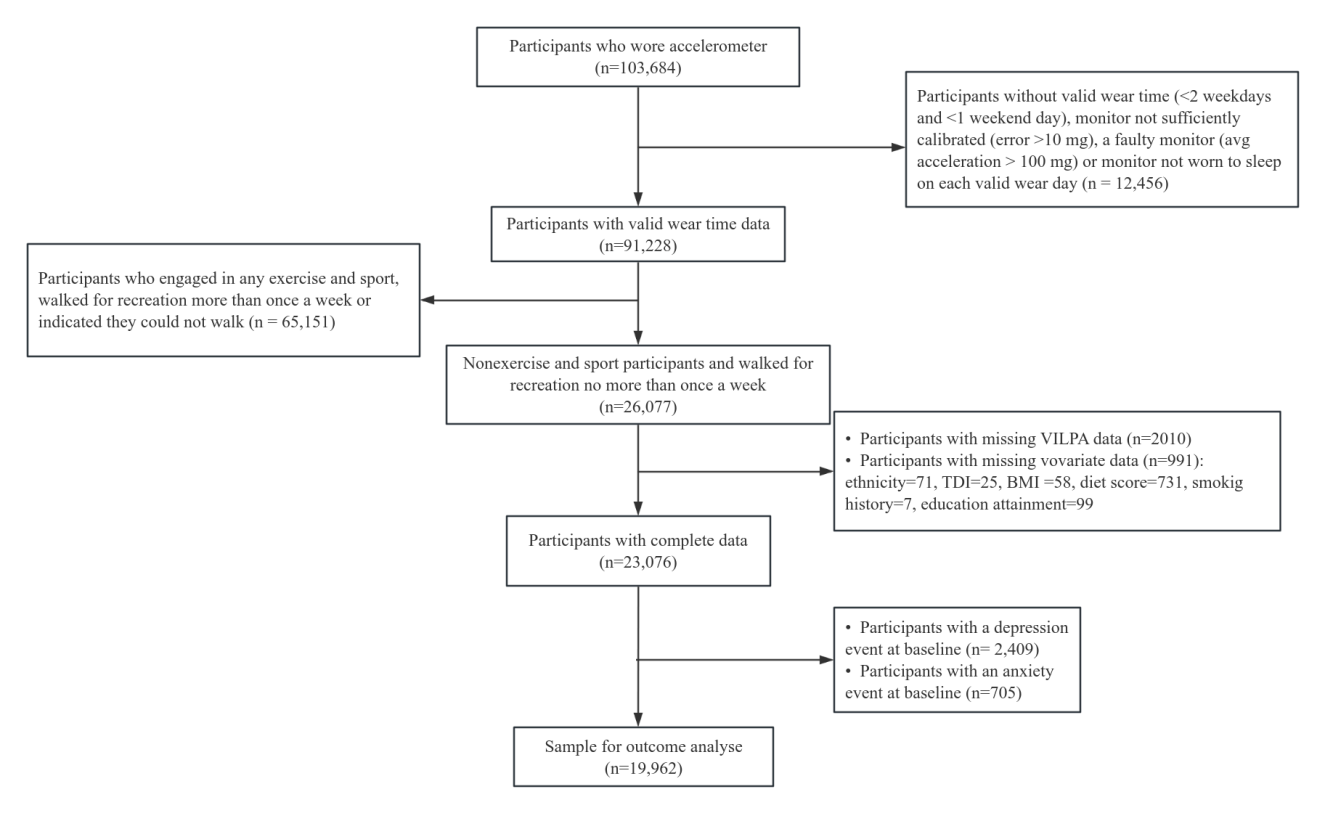

**Supplementary Figure 2. Dose-response association of VILPA duration with risks of depression and anxiety after further adjusting for accelerometer wear season**

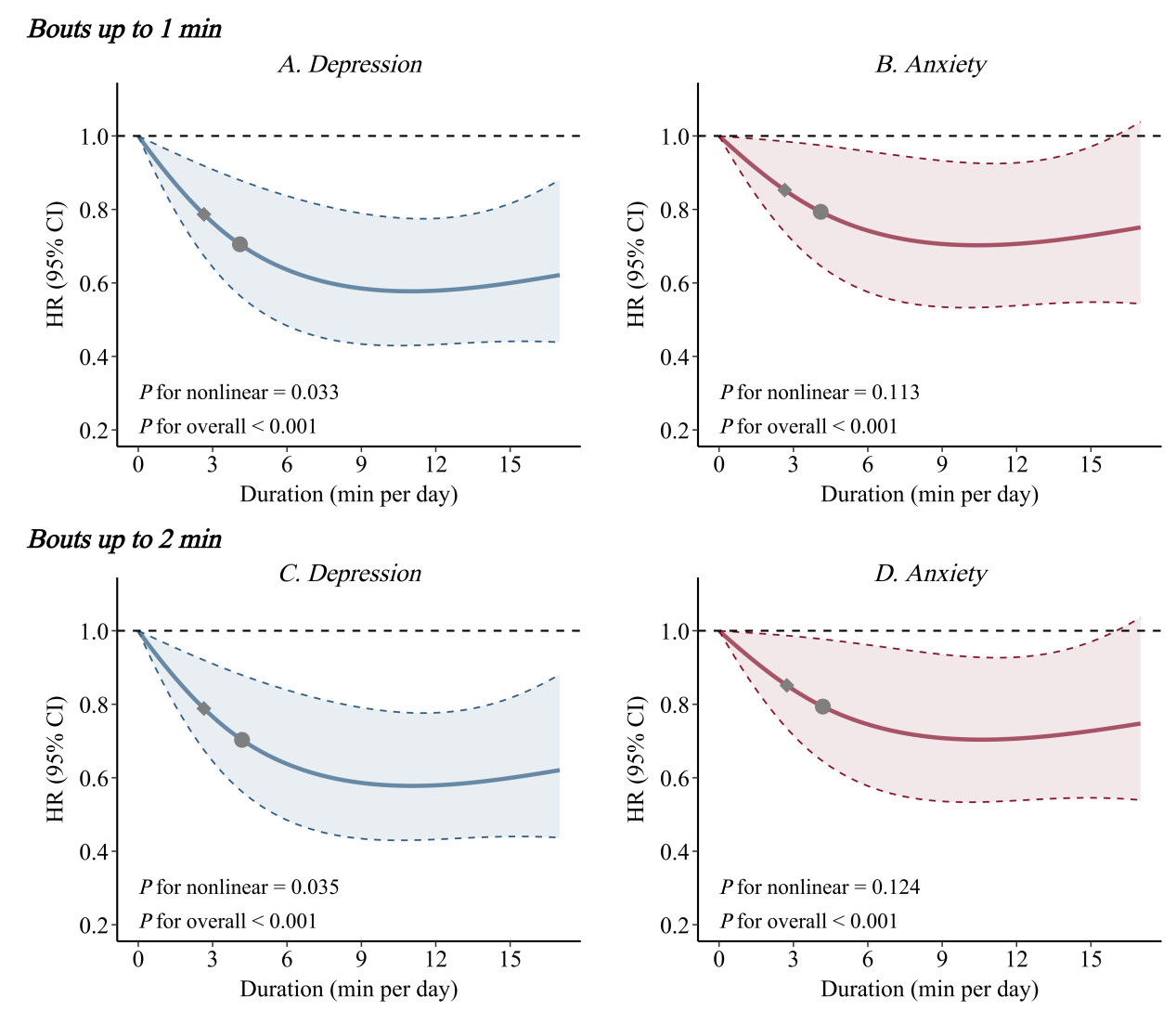

*VILPA*, vigorous intermittent lifestyle physical activity; *HR,* Hazard ration; *CI,* confidence interval.

Panels A and C show depression HRs associated with increasing daily duration of VILPA, for bouts of VILPA up to 1 min (A) and 2 min (C) in duration. Panels B and D show corresponding associations for anxiety. Panel A and C: Depression: n= 19,962; events=469. Panel B and D: Anxiety: n= 19,962; events=536. Diamond, minimal dose, as indicated by the ED_50_ statistic which estimates the daily duration of VILPA associated with 50% of the optimal risk reduction. Circle, HR associated with the median VILPA. The VILPA duration median values were calculated in the sample excluding participants with zero VILPA. Analyses adjusted for age, sex, ethnicity, BMI, educational attainment, TDI, smoking history, drinking status, dietary pattern, and self-reported medication use (diabetes, hypertension, or hypercholesterolemia). Additional covariates included accelerometer estimated sleep duration, PA energy expenditure volume from non-VILPA, daily duration of VPA bouts lasting >1 minute (for analyses of VILPA bouts ≤1 minute) or >2 minutes (for analyses of VILPA bouts ≤2 minutes), and the season of wear (spring, summer, autumn, and winter).

**Supplementary Figure 3. Dose-response association of VILPA duration with risks of depression and anxiety after excluding prevalent cancer at baseline**

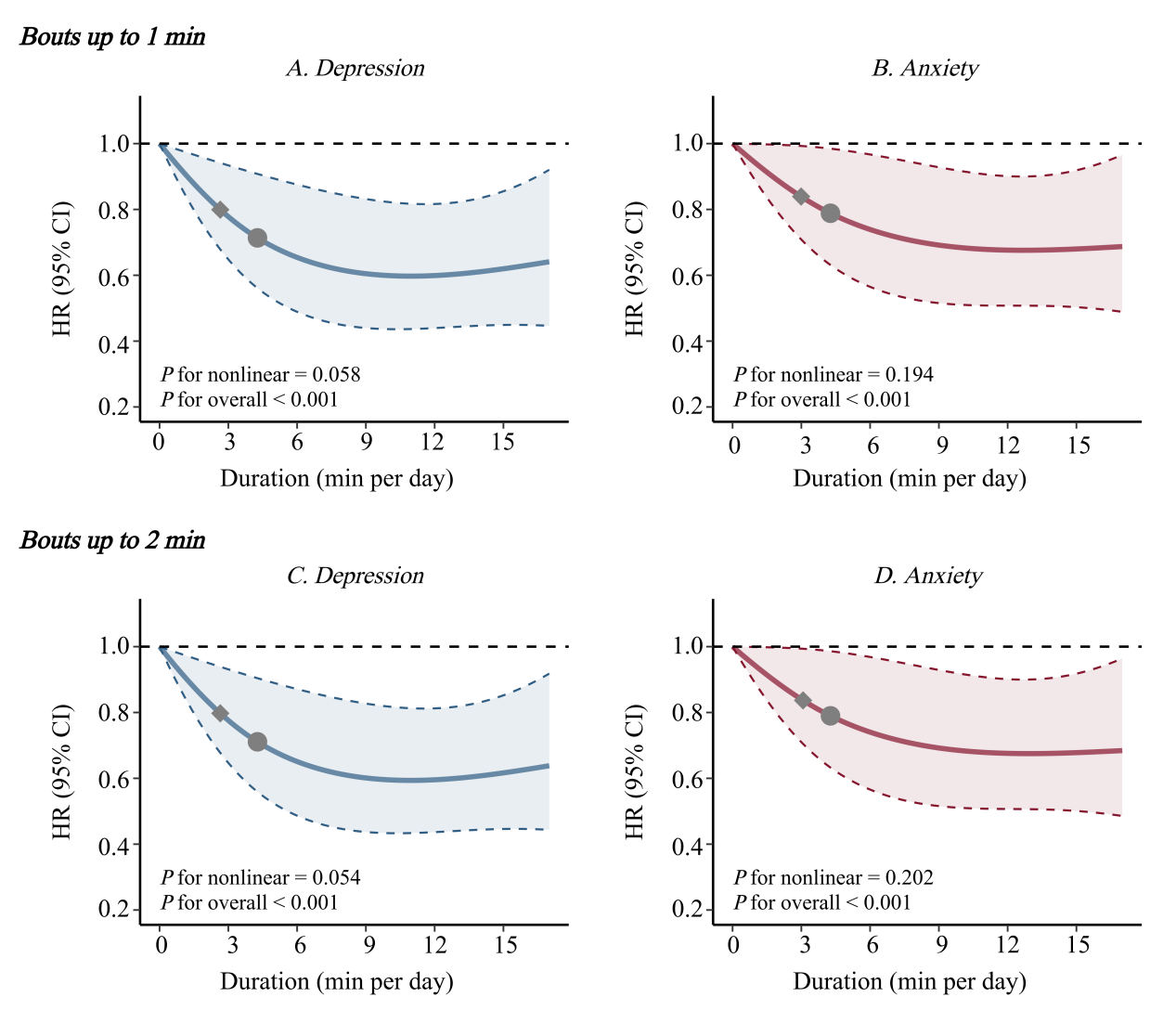

*VILPA*, vigorous intermittent lifestyle physical activity; *HR,* Hazard ration; *CI,* confidence interval.

Panels A and C show depression HRs associated with increasing daily duration of VILPA, for bouts of VILPA up to 1 min (A) and 2 min (C) in duration. Panels B and D show corresponding associations for anxiety. Panel A and C: Depression: n=18,221; events=417. Panel B and D: Anxiety: n=18,221; events=482.Diamond, minimal dose, as indicated by the ED_50_ statistic which estimates the daily duration of VILPA associated with 50% of the optimal risk reduction. Circle, HR associated with the median VILPA. The VILPA duration median values were calculated in the sample excluding participants with zero VILPA. Analyses adjusted for age, sex, ethnicity, BMI, educational attainment, TDI, smoking history, drinking status, dietary pattern, and self-reported medication use (diabetes, hypertension, or hypercholesterolemia). Additional covariates included accelerometer estimated sleep duration, PA energy expenditure volume from non-VILPA, and daily duration of VPA bouts lasting >1 minute (for analyses of VILPA bouts ≤1 minute) or >2 minutes (for analyses of VILPA bouts ≤2 minutes).

**Supplementary Figure 4. Dose-response association of VILPA duration with risks of depression and anxiety after excluding prevalent CVD at baseline**

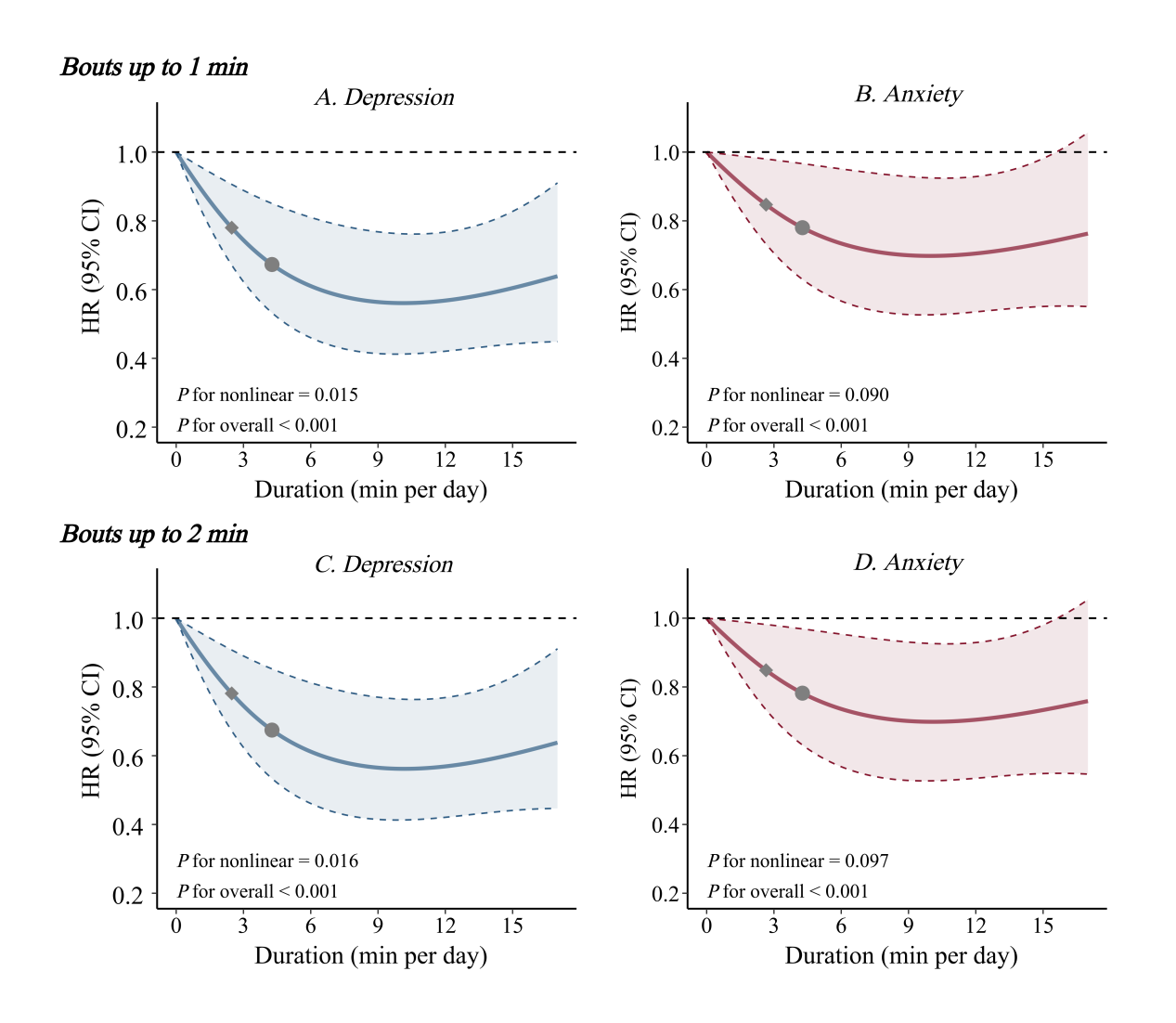

*VILPA*, vigorous intermittent lifestyle physical activity; *HR,* Hazard ration; *CI,* confidence interval.

Panels A and C show depression HRs associated with increasing daily duration of VILPA, for bouts of VILPA up to 1 min (A) and 2 min (C) in duration. Panels B and D show corresponding associations for anxiety. Panel A and C: Depression: n= 19,061; events=440. Panel B and D: Anxiety: n= 19,061; events=518. Diamond, minimal dose, as indicated by the ED_50_ statistic which estimates the daily duration of VILPA associated with 50% of the optimal risk reduction. Circle, HR associated with the median VILPA. The VILPA duration median values were calculated in the sample excluding participants with zero VILPA. Analyses adjusted for age, sex, ethnicity, BMI, educational attainment, TDI, smoking history, drinking status, dietary pattern, and self-reported medication use (diabetes, hypertension, or hypercholesterolemia). Additional covariates included accelerometer estimated sleep duration, PA energy expenditure volume from non-VILPA, and daily duration of VPA bouts lasting >1 minute (for analyses of VILPA bouts ≤1 minute) or >2 minutes (for analyses of VILPA bouts ≤2 minutes).

**Supplementary Figure 5. Dose-response association of VILPA duration with risks of depression and anxiety after excluding outcome diagnosed within 2 years since baseline**

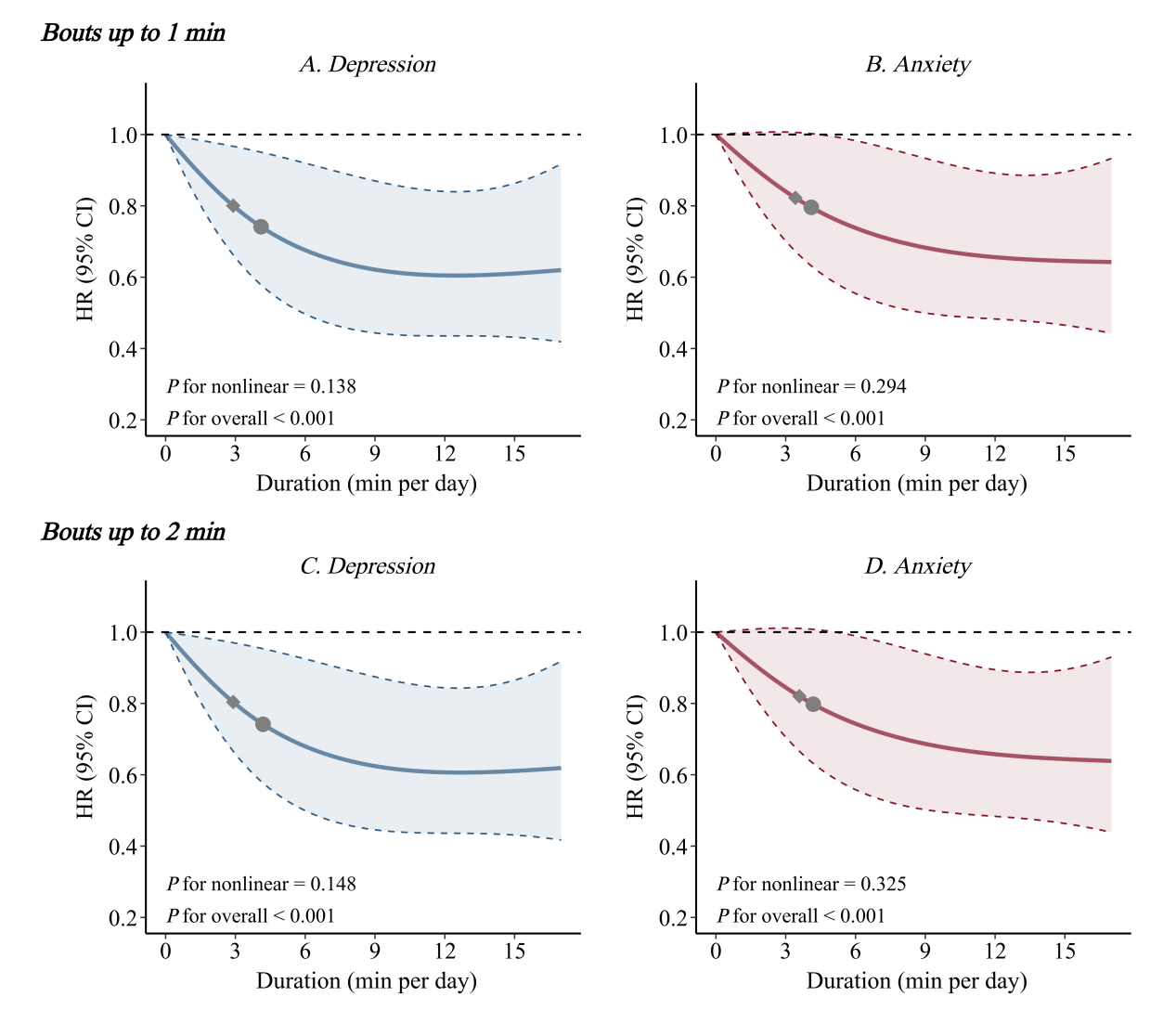
*VILPA*, vigorous intermittent lifestyle physical activity; *HR,* Hazard ration; *CI,* confidence interval.

Panels A and C show depression HRs associated with increasing daily duration of VILPA, for bouts of VILPA up to 1 min (A) and 2 min (C) in duration. Panels B and D show corresponding associations for anxiety. Panel A and C: Depression: n= 19,863; events=370. Panel B and D: Anxiety: n= 19,850; events=424. Diamond, minimal dose, as indicated by the ED_50_ statistic which estimates the daily duration of VILPA associated with 50% of the optimal risk reduction. Circle, HR associated with the median VILPA. The VILPA duration median values were calculated in the sample excluding participants with zero VILPA. Analyses adjusted for age, sex, ethnicity, BMI, educational attainment, TDI, smoking history, drinking status, dietary pattern, and self-reported medication use (diabetes, hypertension, or hypercholesterolemia). Additional covariates included accelerometer estimated sleep duration, PA energy expenditure volume from non-VILPA, and daily duration of VPA bouts lasting >1 minute (for analyses of VILPA bouts ≤1 minute) or >2 minutes (for analyses of VILPA bouts ≤2 minutes).

**Supplementary Figure 6. Dose-response association of VILPA duration with risks of depression and anxiety after excluding participants with poor health**

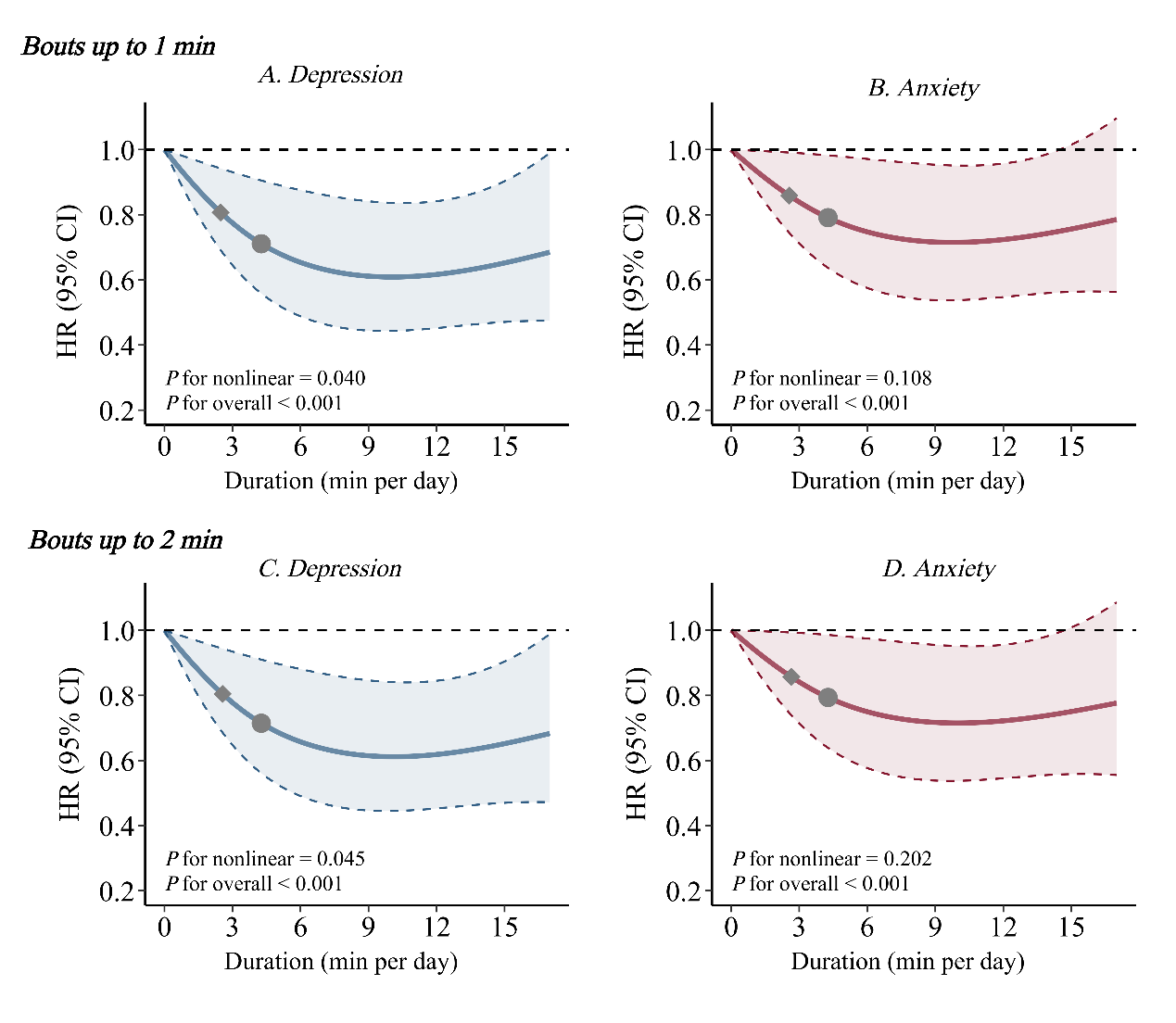

*VILPA*, vigorous intermittent lifestyle physical activity; *HR,* Hazard ration; *CI,* confidence interval.

Panels A and C show depression HRs associated with increasing daily duration of VILPA, for bouts of VILPA up to 1 min (A) and 2 min (C) in duration. Panels B and D show corresponding associations for anxiety. Panel A and C: Depression: n= 19,361; events=412. Panel B and D: Anxiety: n= 19,361; events=507. Diamond, minimal dose, as indicated by the ED_50_ statistic which estimates the daily duration of VILPA associated with 50% of the optimal risk reduction. Circle, HR associated with the median VILPA. The VILPA duration median values were calculated in the sample excluding participants with zero VILPA. Analyses adjusted for age, sex, ethnicity, BMI, educational attainment, TDI, smoking history, drinking status, dietary pattern, and self-reported medication use (diabetes, hypertension, or hypercholesterolemia). Additional covariates included accelerometer estimated sleep duration, PA energy expenditure volume from non-VILPA, and daily duration of VPA bouts lasting >1 minute (for analyses of VILPA bouts ≤1 minute) or >2 minutes (for analyses of VILPA bouts ≤2 minutes).

**Supplementary Figure 7. Dose-response association of VILPA duration with risks of depression and anxiety after excluding participants using antidepressant or antipsychotic medications**

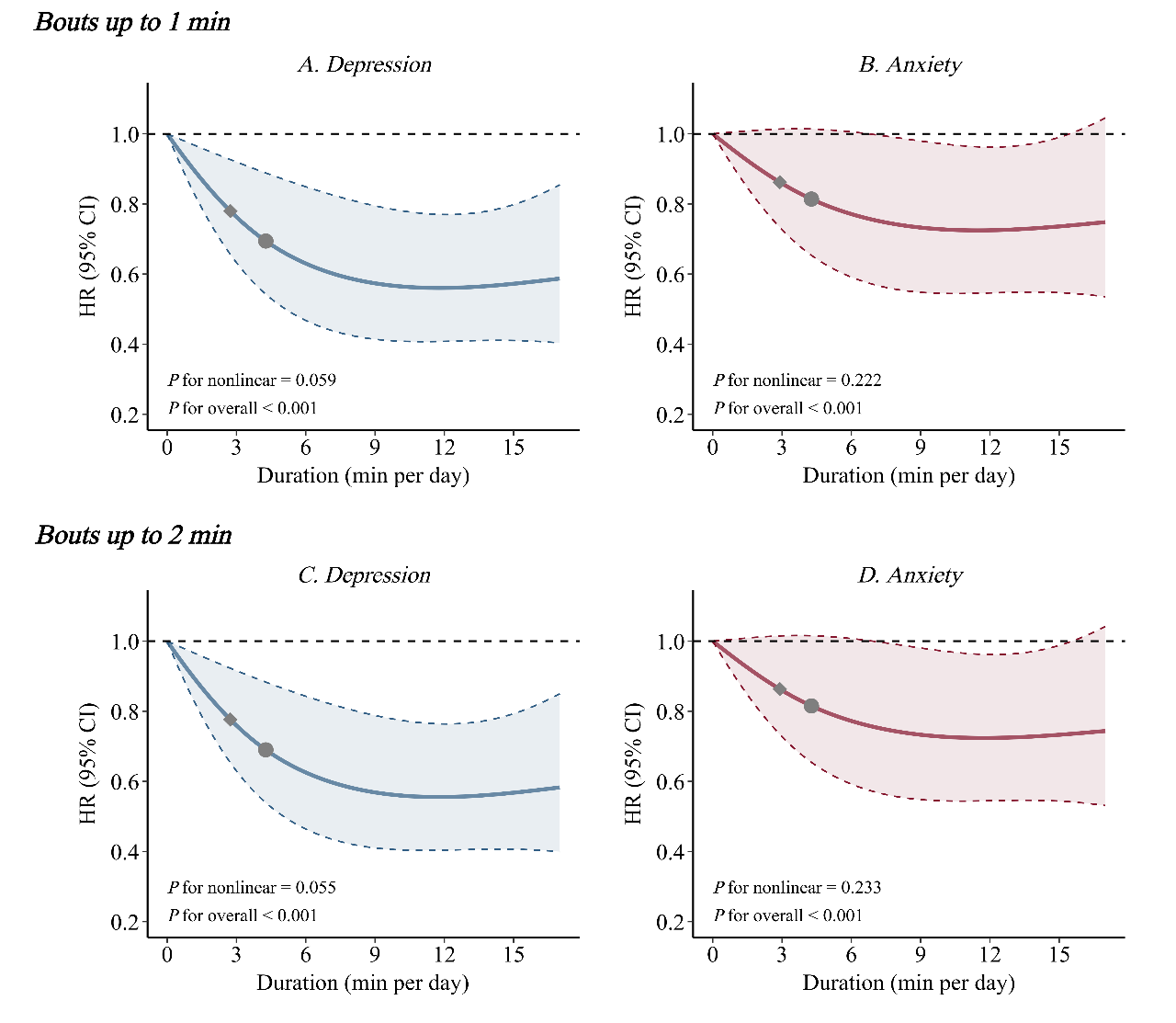
*VILPA*, vigorous intermittent lifestyle physical activity; *HR,* Hazard ration; *CI,* confidence interval.

Panels A and C show depression HRs associated with increasing daily duration of VILPA, for bouts of VILPA up to 1 min (A) and 2 min (C) in duration. Panels B and D show corresponding associations for anxiety. Panel A and C: Depression: n= 19,422; events=396. Panel B and D: Anxiety: n= 19,422; events=492. Diamond, minimal dose, as indicated by the ED_50_ statistic which estimates the daily duration of VILPA associated with 50% of the optimal risk reduction. Circle, HR associated with the median VILPA. The VILPA duration median values were calculated in the sample excluding participants with zero VILPA. Analyses adjusted for age, sex, ethnicity, BMI, educational attainment, TDI, smoking history, drinking status, dietary pattern, and self-reported medication use (diabetes, hypertension, or hypercholesterolemia). Additional covariates included accelerometer estimated sleep duration, PA energy expenditure volume from non-VILPA, and daily duration of VPA bouts lasting >1 minute (for analyses of VILPA bouts ≤1 minute) or >2 minutes (for analyses of VILPA bouts ≤2 minutes).

**Supplementary Figure 8. Dose-response association of VILPA duration with risks of depression and anxiety after excluding participants with VILPA = 0 min /day**

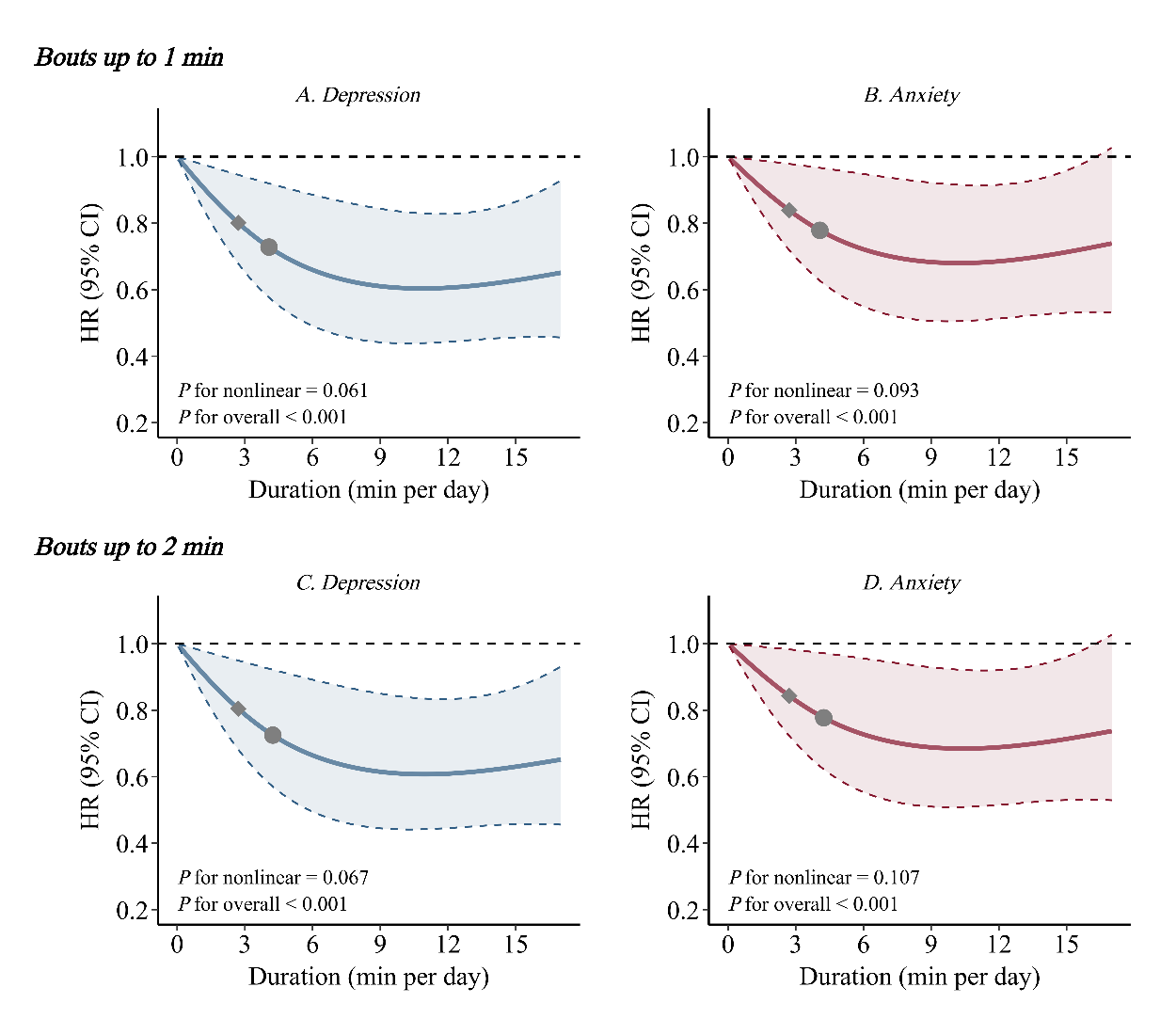
*VILPA*, vigorous intermittent lifestyle physical activity; *HR,* Hazard ration; *CI,* confidence interval.

Panels A and C show depression HRs associated with increasing daily duration of VILPA, for bouts of VILPA up to 1 min (A) and 2 min (C) in duration. Panels B and D show corresponding associations for anxiety. Panel A: n= 18,885; events=432. Panel B: n= 18,885; events=500. Panel A: n= 18,900; events=432. Panel B: n= 18,885; events=500.Diamond, minimal dose, as indicated by the ED_50_ statistic which estimates the daily duration of VILPA associated with 50% of the optimal risk reduction. Circle, HR associated with the median VILPA. The VILPA duration median values were calculated in the sample excluding participants with zero VILPA. Analyses adjusted for age, sex, ethnicity, BMI, educational attainment, TDI, smoking history, drinking status, dietary pattern, and self-reported medication use (diabetes, hypertension, or hypercholesterolemia). Additional covariates included accelerometer estimated sleep duration, PA energy expenditure volume from non-VILPA, and daily duration of VPA bouts lasting >1 minute (for analyses of VILPA bouts ≤1 minute) or >2 minutes (for analyses of VILPA bouts ≤2 minutes).

**Supplementary Figure 9. Dose-response association of VILPA duration with risks of depression and anxiety after excluding participants with baseline depressive and anxiety symptoms**

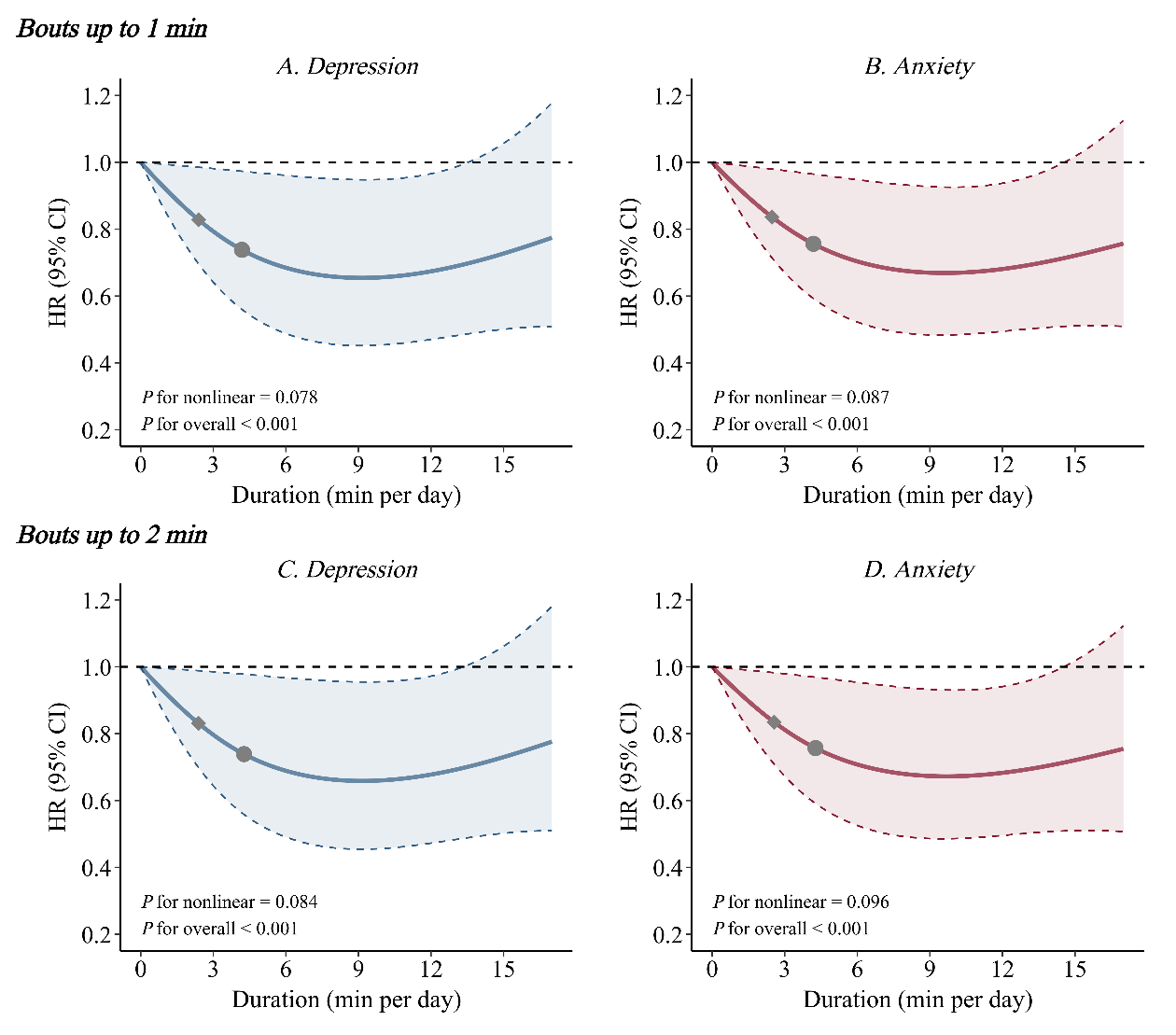

*VILPA*, vigorous intermittent lifestyle physical activity; *HR,* Hazard ration; *CI,* confidence interval.

Panels A and C show depression HRs associated with increasing daily duration of VILPA, for bouts of VILPA up to 1 min (A) and 2 min (C) in duration. Panels B and D show corresponding associations for anxiety. Panel A and C: Depression: n= 16,618; events=307. Panel B and D: Anxiety: n= 16,618; events=393. Diamond, minimal dose, as indicated by the ED_50_ statistic which estimates the daily duration of VILPA associated with 50% of the optimal risk reduction. Circle, HR associated with the median VILPA. The VILPA duration median values were calculated in the sample excluding participants with zero VILPA. Analyses adjusted for age, sex, ethnicity, BMI, educational attainment, TDI, smoking history, drinking status, dietary pattern, and self-reported medication use (diabetes, hypertension, or hypercholesterolemia). Additional covariates included accelerometer estimated sleep duration, PA energy expenditure volume from non-VILPA, daily duration of VPA bouts lasting >1 minute (for analyses of VILPA bouts ≤1 minute) or >2 minutes (for analyses of VILPA bouts ≤2 minutes). Baseline depressive and anxiety symptoms were assessed using the depression and anxiety subscales of the Patient Health Questionnaire-4 (PHQ-4; score range 0–6). Probable depressive or anxiety symptoms were defined as PHQ-4 subscale scores ≥3. In analyses of depression, models were further adjusted for baseline depressive symptoms, whereas in analyses of anxiety, models were further adjusted for baseline anxiety symptoms.

**Supplementary Figure 10. Association of VILPA duration with risks of depression and anxiety using the minimal dose as the referent point**

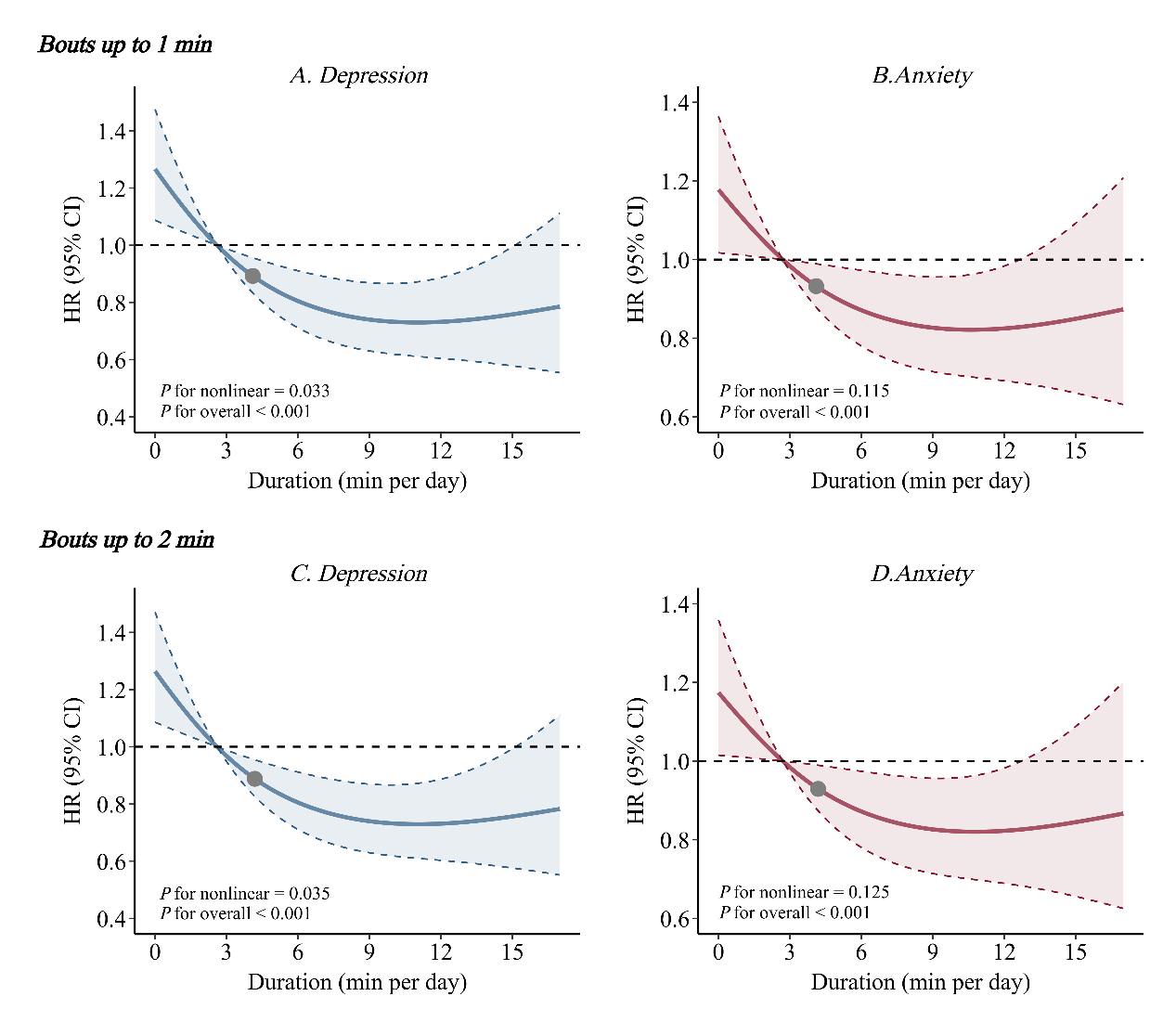

*VILPA*, vigorous intermittent lifestyle physical activity; *HR,* Hazard ration; *CI,* confidence interval.

Panels A and C show depression HRs associated with increasing daily duration of VILPA, for bouts of VILPA up to 1 min (A) and 2 min (C) in duration. Panels B and D show corresponding associations for anxiety. Circle, HR associated with the median VILPA. The VILPA duration median values were calculated in the sample excluding participants with zero VILPA. Analyses adjusted for age, sex, ethnicity, BMI, educational attainment, TDI, smoking history, drinking status, dietary pattern, and self-reported medication use (diabetes, hypertension, or hypercholesterolemia). Additional covariates included accelerometer estimated sleep duration, PA energy expenditure volume from non-VILPA, and daily duration of VPA bouts lasting >1 minute (for analyses of VILPA bouts ≤1 minute) or >2 minutes (for analyses of VILPA bouts ≤2 minutes).

**Supplementary Figure 11. Association of VILPA duration with risks of depression and anxiety after treating death as a competing event**

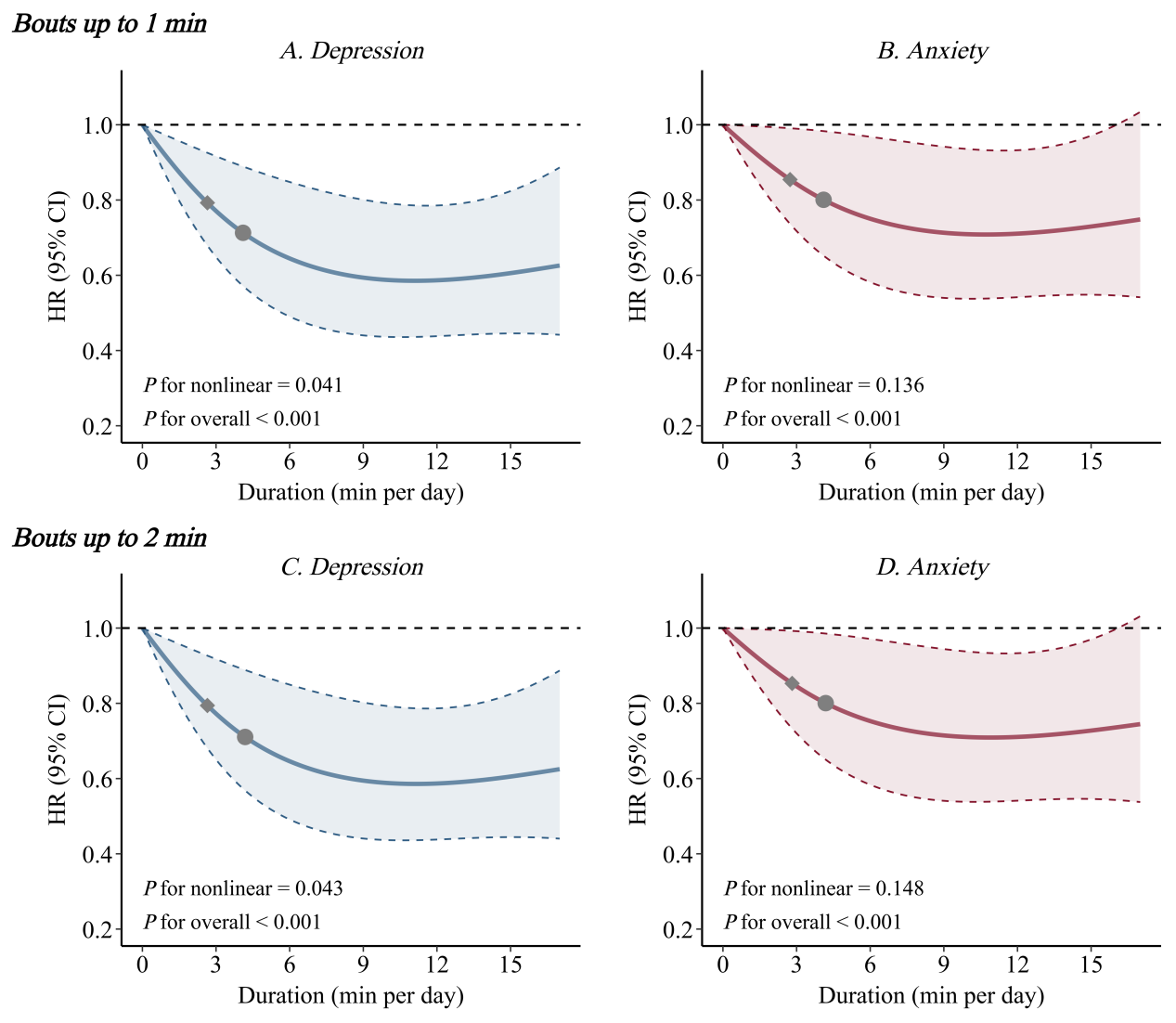

*VILPA*, vigorous intermittent lifestyle physical activity; *HR,* Hazard ration; *CI,* confidence interval.

Panels A and C show depression HRs associated with increasing daily duration of VILPA, for bouts of VILPA up to 1 min (A) and 2 min (C) in duration. Panels B and D show corresponding associations for anxiety. Diamond, minimal dose, as indicated by the ED_50_ statistic which estimates the daily duration of VILPA associated with 50% of the optimal risk reduction. Circle, HR associated with the median VILPA. The VILPA duration median values were calculated in the sample excluding participants with zero VILPA. Analyses adjusted for age, sex, ethnicity, BMI, educational attainment, TDI, smoking history, drinking status, dietary pattern, and self-reported medication use (diabetes, hypertension, or hypercholesterolemia). Additional covariates included accelerometer estimated sleep duration, PA energy expenditure volume from non-VILPA, and daily duration of VPA bouts lasting >1 minute (for analyses of VILPA bouts ≤1 minute) or >2 minutes (for analyses of VILPA bouts ≤2 minutes). Fine-Gray subdistribution hazards to account for competing mortality risk.

**Supplementary Methods: Wearable physical activity intensity and posture classification**

Incidental physical activity was classified using a validated two-stage random forest activity classifier that first classifies each 10 s window (epoch) as sedentary (lying or sitting still), stationary plus (active sitting, standing still, active standing), walking, or running (**Diagram A**)^1-3^. These activities were then classified into one of four activities including: sedentary, light, moderate, and vigorous. Walking activities (gardening, active commuting, etc) were classified by normalized gravitational units (g) where <100 milli g were classified as light intensity (<3 METs), ≥100 milli g and <400 milli g were considered moderate intensity physical activity (≥3 to <6 METs), and ≥400 milli g were considered vigorous-intensity PA (≥6 METs)^3^. All windows classified as running/high energetic activity were classified as vigorous-intensity physical activity (≥ 6 METs)^3-5^.

**Physical Activity Classification Scheme (Diagram A)^5^.**

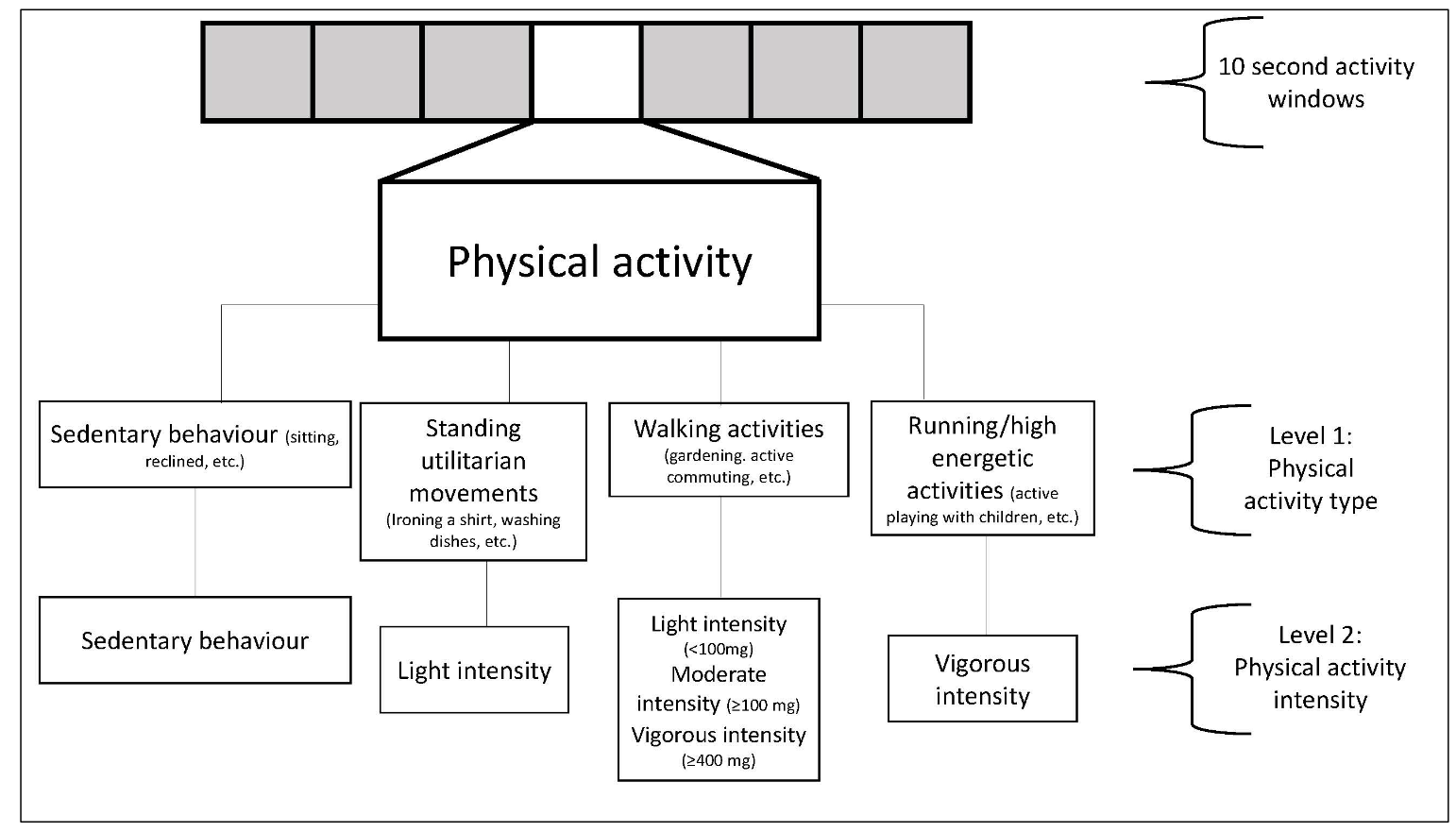

**Physical Activity Classification Performance**

The performance of this physical activity classification scheme was tested in an independent sample of 102 adults from the US^6^ and Australia^7^. This data includes direct observation measurement of 105,767 activity samples from structured and free-living activities (17,627 minutes) which were used to test the robustness and generalizability of the two-stage activity and intensity classifier. The data was collected from participant-worn or researcher-held Go-Pro video recordings. All data was imported into Noldus Observer XT software for continuous video coding. The direct observation coding generated continuous physical activity codes corresponding to the start and finish of each movement. These coded movements were then compared against the accelerometer data using the available time-stamp information. The below table includes the performance metrics across activities. Interobserver reliability was assessed by dual coding. The intraclass correlation coefficient for coding activities was 0.912 (0.866-0.942). The performance in metrics and confusion matrix for activity classification is shown below.

**Classifier Performance Metrics for Intensity in US and Australian Adults^5^**

|  | Sensitivity | Specificity | Precision | F-score | Overall Accuracy | Weighted Kappa | Overall F-score |
| --- | --- | --- | --- | --- | --- | --- | --- |
| Sedentary | 86.5 | 93.7 | 90.5 | 88.5 |  |  |  |
| Light | 71.2 | 89.4 | 55.8 | 62.6 |  |  |  |
| Moderate | 85.4 | 96.6 | 92.7 | 88.9 |  |  |  |
| Vigorous | 95.4 | 99.4 | 94.6 | 95.0 |  |  |  |
|  |  |  |  |  | **84.6** | 0.78 | 83.8 |

Rows= ground truth; columns=predictions; bold=correct classification; all activities were free-living or simulated free-living activities.

**Confusion Matrix for Activity Classification in US and Australian Adults^5^**

|  | Sedentary | Light | Moderate | Vigorous |
| --- | --- | --- | --- | --- |
| Sedentary | **36,904** | 5,232 | 508 | 2 |
| Light | 3,120 | **11,712** | 1,612 | 17 |
| Moderate | 502 | 4,016 | **29,528** | 526 |
| Vigorous | 226 | 17 | 214 | **9,470** |
| Rows= ground truth; columns=predictions; bold=correct classification; all activities were free-living or simulated free-living activities. | | | | |
